# Mapping the Neuroanatomy of Dystonia Using Causal Brain Lesions

**DOI:** 10.64898/2025.12.07.25341373

**Authors:** Daniel T. Corp, Jaakko Kungshamn, Elizabeth G. Ellis, Maximilian U. Friedrich, Kalle J. Niemi, Christopher J. Greenwood, Jordan Morrison-Ham, Shiro Horisawa, Martin M. Reich, Hyder A. Jinnah, Andreas Horn, Michael D. Fox, Juho Joutsa

**Affiliations:** Cognitive Neuroscience Unit, School of Psychology, Deakin University, Burwood, Australia; Turku Brain and Mind Center, Clinical Neurosciences, University of Turku, Turku, Finland; Fondazione Policlinico Universitario Campus Bio-Medico, Via Alvaro del Portillo 200, 00128 Roma, Italy; Center for Brain Circuit Therapeutics, Departments of Neurology, Psychiatry, and Radiology, Brigham & Women’s Hospital, Boston; Department of Neurology, University Hospital Ulm, 89081 Ulm, Germany; Deakin University, SEED Lifespan Strategic Research Centre, School of Psychology, Faculty of Health, Geelong, Australia; Murdoch Children’s Research Institute, Centre for Adolescent Health, Melbourne, Australia; The University of Melbourne, Department of Paediatrics, Royal Children’s Hospital, Australia; Department of Neurosurgery, Tokyo Women’s Medical University, Tokyo, Japan; Department of Neurology, University Hospital of Würzburg, Josef-Schneider-Straße 11, 97080 Würzburg, Germany; Departments of Neurology, Human Genetics, and Pediatrics, Emory University, Atlanta, GA, United States; Institute for Network Stimulation, Department of Stereotactic and Functional Neurosurgery, University Hospital Cologne, Germany; Neurocenter, Turku University Hospital, Turku, Finland

## Abstract

Dystonia is one of the most prevalent movement disorders, but its neural substrates have remained enigmatic despite decades of research. Brain lesions leading to dystonia offer unique causal inference that could lend insight into the neural networks driving the disorder. Here, we studied the published cases of lesions causing dystonia to a variety of different body parts (n=179) and used lesion network mapping to test whether lesion locations mapped to a common brain network. Specificity was investigated by comparing the findings to 216 lesions causing other movement disorders, and 499 lesions causing non-specific neurological symptoms. While lesions causing dystonia occurred in heterogeneous locations, they localised to a common and specific cortico-basal ganglia-cerebellar network. Stronger connectivity to this dystonia network from a causal brain lesion was associated with a greater extent of dystonia throughout the body. Further, there were significant differences in the brain networks depending on the body parts affected by dystonia, which also demonstrated different somatotopic representations in the cerebellum. Our results suggest that dystonia stems from a cortico-basal ganglia-cerebellar network, with the manifestation in the body depending on the preferential involvement of the hubs of the network, and its somatotopy.

## Introduction

Dystonia is a chronic neurological disorder characterised by sustained and involuntary contractions of the muscles^1^. Despite dystonia being one of the most common movement disorders^3^, the key neural structures or mechanisms are not yet well characterised. Traditionally, dystonia was considered a disorder of the basal ganglia^4,5^, yet an accumulation of neuroimaging, neurophysiological, and brain lesion data over the past 20 years has shown that this view is incomplete^6–9^. Instead, the prevailing view is now that dystonia is a ‘network disorder’, with involvement of multiple communicating brain regions^7,10^. However, there is lack of agreement, especially in the neuroimaging literature, about what the nodes of this network are^7,11^. There is also a high degree of variability across patients in which body part is affected by dystonia and how widespread dystonia is throughout the body. This clinical heterogeneity might be contributing to the lack of uniform findings across studies.

In addition, case-control neuroimaging studies are correlational by nature, not allowing for causal inference from brain abnormalities to symptoms^12,13^. In contrast, the study of focal brain lesions can establish causal relationships between brain damage and neurological symptoms. For example, traditional lesion studies have demonstrated that lesions impairing memory localise to the medial temporal lobes, and lesions impairing speech production to the left frontal lobe (Broca’s area)^14–16^. However, many neurological symptoms can result from lesions in different brain locations^17^. A recent study investigating all published cases of lesion-induced dystonia (n=359) showed that, although the basal ganglia was the most common lesion location, there was not a unifying anatomical structure underlying dystonia, and in fact, most of the lesions occurred outside the basal ganglia^8^. Further, while there were relationships between lesion location and the body parts affected by dystonia, most lesions resulting in dystonia of a specific body part did not intersect just one neural structure. For example, hand dystonia was associated with lesions to the thalamus, yet only 33.5% of cases with hand dystonia had a thalamic lesion.

Lesion network mapping is a novel technique combining lesion locations with brain connectomes (‘wiring diagrams of the human brain’^17^), which extends lesion localisation from anatomical regions to brain networks^17,18^. This approach has been used to successfully map lesions causing numerous different neurological and psychiatric symptoms onto brain networks^19–23^. Previously, we used this technique to show that 25 lesions causing cervical dystonia (CD) mapped to a common brain network^24^. However, whether lesions causing different types of dystonia share a common brain network, or map to distinct brain networks, remains unknown. Here we use lesion network mapping to study 179 published cases of lesion locations causing dystonia across the different body parts.

## Methods

### Case selection

Cases of lesion-induced dystonia were identified from a systematic search of Pubmed and Embase including cases up to July 2022. See Supplementary file 1 for search terms and Supplementary File 2 for PRISMA flowchart. Our prior inclusion/exclusion criteria for lesion-induced dystonia are provided in detail in Corp et al.^8^. Briefly, inclusion criteria were a published report describing human patient/s with lesion-induced dystonia, including a neurological examination where the authors of the report concluded that the dystonia was caused by a lesion. The same criteria were used here, yet with five additional exclusion criteria given that in this study lesions needed to be traced for lesion network mapping analyses: 1) lesions in children aged <10, given that in these cases the brain anatomy does not sufficiently resemble the standard adult brain^24^; 2) very large or diffuse lesions with poorly identifiable borders, spanning multiple sections of functionally heterogeneous brain tissue^25,26^; 3) lesions outside of the brain (e.g., spinal cord); 4) no figure of the lesion(s); 5) poor scan or poor quality figure of the lesion in the published report precluding accurate delineation of the lesion. Data from published reports were independently extracted by D.C. and J.K., with patients’ symptoms and characteristics classified as previously described^8^. Any discrepancies in the extracted data were resolved by J.J..

### Lesion network mapping across all dystonias

The network of brain regions functionally connected to each lesion location (Figure 1) was identified by performing lesion network mapping as previously described^19,24^. First, lesions from published images were traced by hand onto a MNI152 2mm standardised brain atlas using FSLeyes software^27^. Lesion tracings were performed by D.C., J.K., and M.Fr., cross-checked by another of these three authors, and discrepancies resolved by J.J.. Functional connectivity maps were created for each lesion (in Lead DBS software [https://www.lead-dbs.org])^28^ using a standard seed-based approach, leveraging rs-fMRI data from a normative dataset of 1000 healthy young adults^29,30^, resulting in a connectivity t-map for each lesion (Figure 2A).

**Figure 1.**
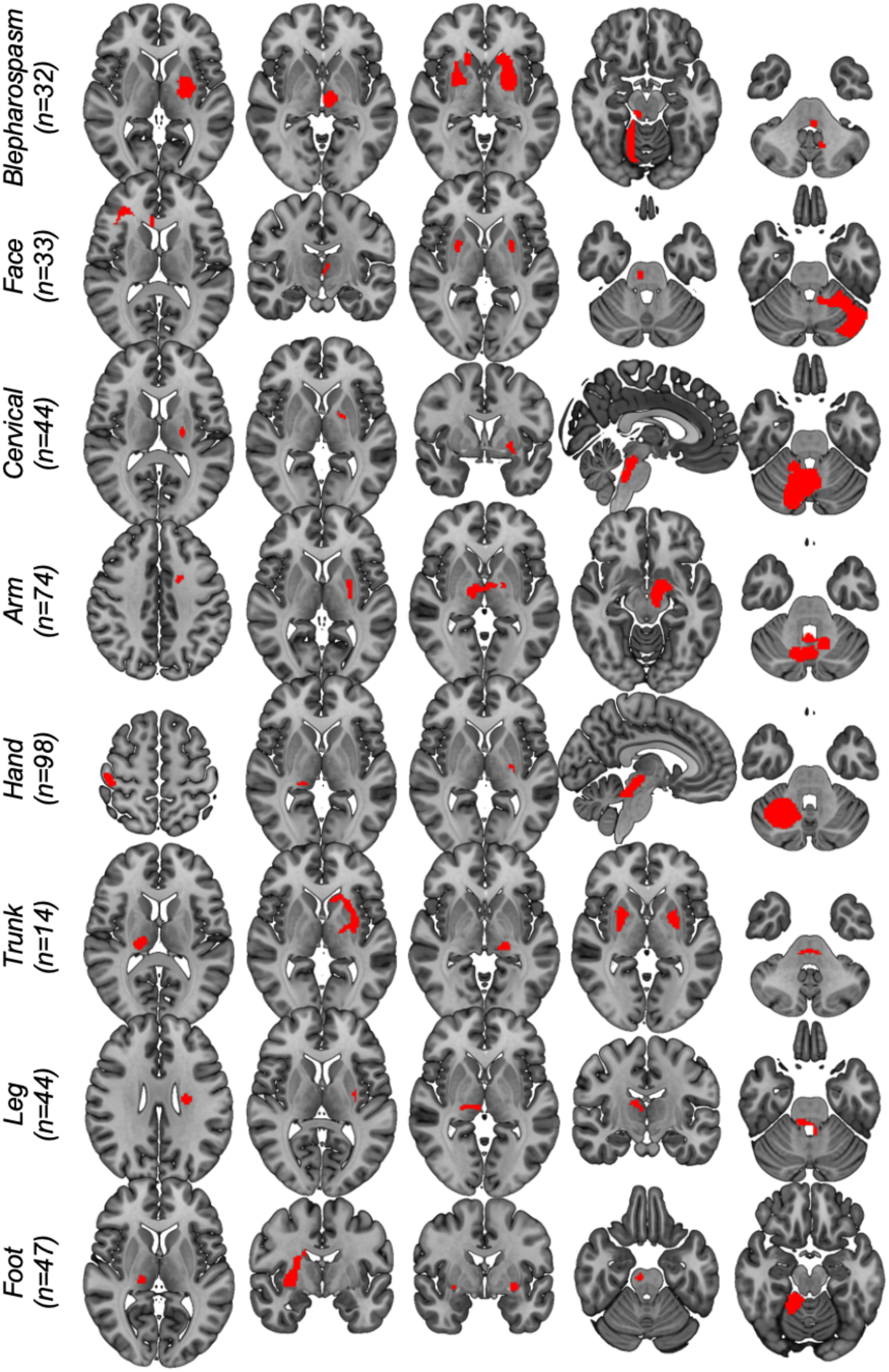
Lesions causing dystonia. Example lesions causing dystonia from the total sample of 179. For each body part affected by dystonia, lesions occurred in numerous different brain regions.

**Figure 2.**
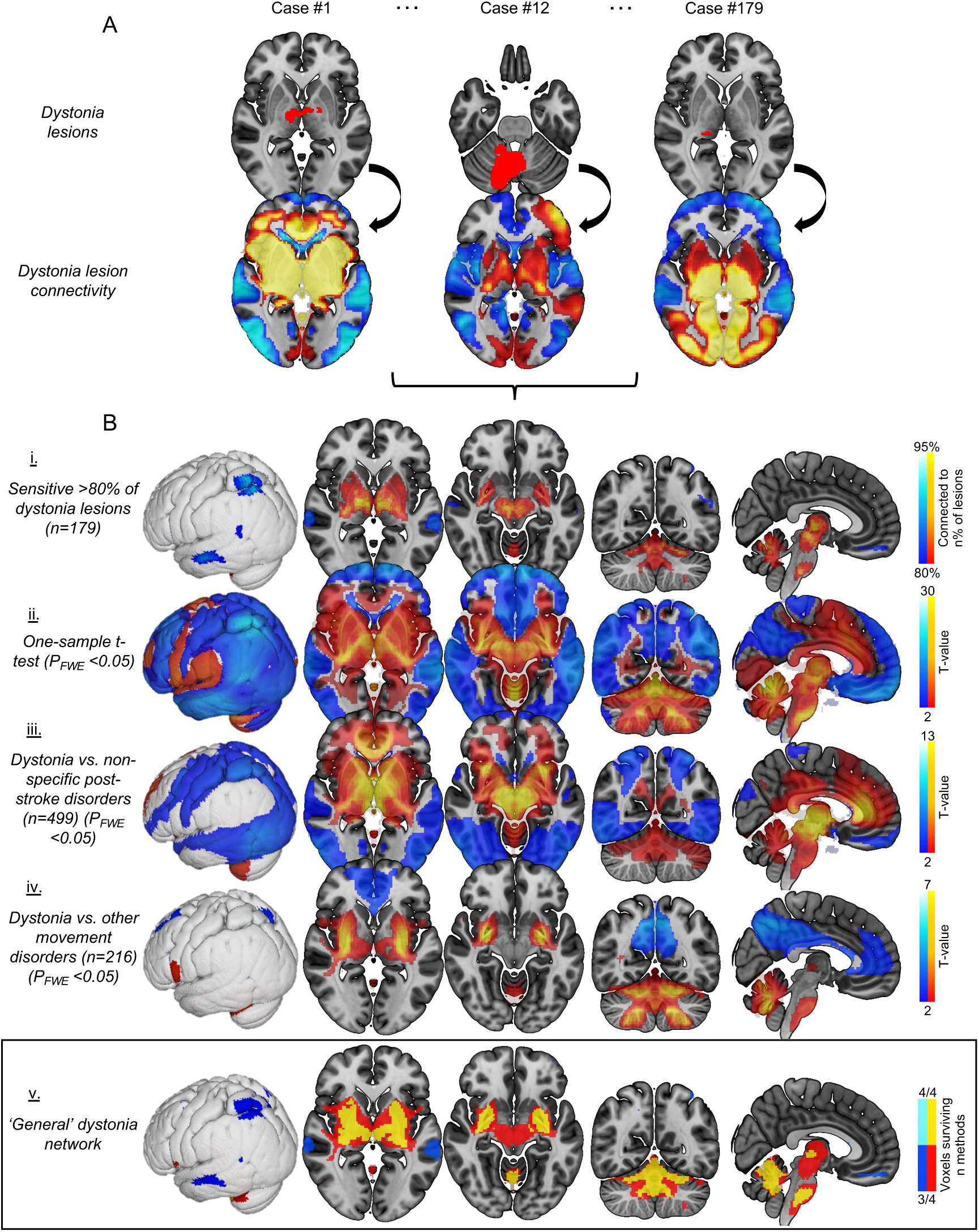
General dystonia network. Lesions causing dystonia were traced to a standard adult brain and functional connectivity between each lesion and the rest of the brain was computed using a normative dataset of rs-fMRI scans from 1000 healthy individuals (Figure A). These t-maps were then used in a series of analyses to determine the neural structures most connected to causal lesions (Figure B, i-iv). Across these analyses, a set of key brain regions were consistently implicated. We defined the ‘general dystonia network’ as the voxels surviving ≥3 of these four statistical tests (Figure B, v). T-values are shown for visual purposes in rows ii-iv, masked to regions p<0.05 FWE corrected.

To identify the brain networks connected to lesion locations causing dystonia, we performed four analyses using different techniques and control groups (Figure 2B). This was done to find the neural structures that were robust to statistical methods and comparison groups. First, maps were thresholded at t-value of ≥7 corresponding to whole brain family-wise error (FWE) corrected P<10^-6^ (as in our prior study focusing on CD^24^), binarised (positive and negative connectivity maps run separately as they may have different biological interpretation), and then overlapped to identify voxels connected to ≥80% of causal lesions. Second, a one-sample t-test was conducted to identify voxels of lesion connectivity significantly different from zero using FSL randomise^27,31^. Third, a two-sample t-test was conducted to identify voxels with significantly different connectivity from dystonia lesion locations compared to those of a control dataset of 499 stroke lesion locations causing a range of neurological symptoms other than dystonia (termed ‘non-specific neurological disorders’) collected from Turku University Hospital medical records (see Supplementary File 3 for cohort information). Fourth, dystonia lesion connectivity was compared to a second control dataset of 216 lesions causing other (non-dystonia) movement disorders from prior lesion network mapping studies. This was the same control dataset as used in our previous analysis of lesion locations in dystonia^8^, including lesions causing parkinsonism (29 cases), alien limb syndrome (50), akinetic mutism (28), hemichorea-hemiballismus (29), asterixis (30), freezing of gait (14), and Holmes tremor (36)^20,21,32–35^. Given the multiple disorders within this control group, a general linear model with planned contrasts (equal weights for each control disorder) was conducted using FSL randomise^21,25^. The final brain network associated with dystonia (‘general dystonia network’) was then derived from a conjunction analysis identifying the voxels surviving ≥3 of these four statistical tests. All FSL randomise analyses in the paper used threshold-free cluster enhancement (TFCE) with 5000 permutations, with whole brain FWE corrected P-values <0.05 considered significant^25,36^.

### Network connectivity and extent of dystonia in the body

Next, we investigated whether stronger lesion connectivity to this dystonia brain network could predict a greater extent of dystonia in the body. To do this, for each case’s lesion network map, the mean absolute functional connectivity strength (t-value) of voxels that overlapped with the general dystonia network was computed, and this value was then associated with: 1) the number of body parts affected by dystonia, using Spearman rank correlation; and 2) dystonia body distribution (focal, segmental, multifocal, generalised, or hemidystonia), using multiple linear regression, with post-hoc pairwise tests where regression showed significance. Each case’s dystonia body distribution was categorised as in our previous study on lesion-induced dystonia^8^, aligning with consensus statements on the phenomenology and classification of dystonia^1,2^.

To test whether the extent of dystonia in the body was specifically associated with lesion connectivity to the dystonia network, two additional analyses were conducted. First, we investigated whether the mean absolute t-value of voxels across the *whole brain* predicted the number of body parts affected or dystonia body distribution (as opposed to the mean absolute t-value of voxels specifically overlapping with the general dystonia network, performed above). Second, we assessed whether *lesion size* (rather than lesion connectivity, as above) predicted the same outcomes.

To confirm these findings, we also tested whether the *number of voxels* (as opposed to connectivity strength, above) with a connectivity value of t≥7 (same threshold as in overlap analysis, above) that overlapped with the general dystonia network predicted the extent of dystonia in the body. These analyses were conducted using StataSE software (version 15.1, StataCorp LLC: College Station, TX).

### Network connectivity and dystonia symptom latency

As the onset of dystonia following the lesion is variable, we then investigated whether lesion connectivity was associated with symptom latency in cases that reported latency information (n=108), using the same approach as above (‘Network connectivity and extent of dystonia in the body’). The mean absolute t-value of voxels overlapping with the general dystonia network was Spearman correlated with symptom latency (latency expressed in weeks from the lesion until dystonia symptom onset, as in Corp et al.^8^). In addition, to identify brain regions specifically associated with latency, we conducted a voxel-wise analysis using FSL randomise (TFCE with 5000 permutations) across the whole brain and within the general dystonia network.

### Lesion network mapping by affected body region

In addition to analysing all 179 cases together, we also mapped networks according to the affected body regions (blepharospasm, face [excluding blepharospasm], cervical, arm, hand, trunk, leg, and foot – non-exclusive groups) and distributions (focal, segmental, multifocal, generalised, or hemidystonia)^1,2^. Primary analyses were conducted on the body part affected, and secondary analyses on body distributions, given that lesion locations predicted dystonic body parts but not distributions in our prior study^8^. Networks for each body part and distribution were derived using the same methods used to localise the general dystonia network across all cases, yet thresholded to voxels connected to ≥90% of causal lesions in overlap analyses given the greater sensitivity of these stratified analyses. Voxels surviving ≥3/4 methods were defined as the lesion network for each of these dystonia types.

To test whether brain networks were significantly different between the different types of dystonia, lesion connectivity patterns were compared within dystonias. To do this, we ran two-sample t-tests using FSL randomise as described above, repeated for each of the different dystonia body parts and distributions. Specifically, each group of interest (e.g., CD) was compared to all other dystonia cases without dystonia to that body part/distribution (e.g., all cases without CD). T-tests were used here rather than a single GLM because these body part groups were non-exclusive (i.e., many cases had multiple body parts affected), therefore a GLM would have involved many cases represented across multiple groups. For each body part/distribution, voxels that showed statistical significance, and also fell within their corresponding body part/distribution lesion network (as described above), were presented.

### Somatotopic representation of dystonic body parts

We assessed whether the networks for different dystonic body parts localised to different anatomical sub-regions within the same brain structure^37–40^. To do this, cervical, hand, and foot dystonia networks were overlaid onto a standard adult brain, masked to the cerebellum (MNI structural atlas, binarised at 25% probability), and visually compared to somatotopic representations of the cerebellum previously defined in healthy subjects by Buckner et al.^41^. We focused primarily on the cerebellum here because: 1) this region was strongly implicated across dystonias affecting different body regions; 2) the somatotopy of this structure is well defined in previous literature^41–43^; and 3) the size of the cerebellum is sufficient for investigating somatotopic representation with the spatial resolution of rs-fcMRI ^41–43^. Cervical, hand, and foot body part networks were selected because they provided analogues to the body parts used in Buckner et al.^41^, and they each demonstrated significant findings compared to other dystonia body parts (see Results). Above, voxels surviving ≥3/4 methods were defined as the lesion networks for each of these body parts. Here, as foot dystonia had a relatively small cerebellar network compared to the other two groups, these thresholds were adjusted to enable visualisation of clusters of comparable size (cervical and hand dystonia: voxels surviving 4/4 tests; foot dystonia: voxels surviving ≥2/4 tests. Maps were also overlaid onto the cerebellar atlas within the SUIT (‘a spatially unbiased atlas template of the human cerebellum’) toolbox (in 2mm space to match lesion network maps)^44,45^ to identify the anatomical sub-regions in which the networks fell. To assess whether these different body parts affected by dystonia also had different representations in other brain regions, we visualised these same network maps within the thalamus and putamen, two other commonly localised brain structures in our analyses.

### Visualisation of networks

For each of the identified networks, slices were created using MRIcroGL software (version 14.4) on a MNI152 2009bet 0.5mm brain, and surface maps were created using Mango software (version 4.1) on a MNI152 T1 1mm brain. For the somatotopy analysis, dystonia maps and atlas regions were overlaid on a high-resolution 500µm ex-vivo MNI brain^46^ to aid visualisation (Mango software, version 4.1).

## Results

### Lesions causing dystonia

The systematic search identified 179 cases of lesion-induced dystonia where the lesion could be clearly delineated. A description of all included cases is provided in Supplementary File 4. Lesions occurred across multiple heterogeneous brain regions, even when grouped by dystonia type (Figure 1).

### Lesion network mapping across all dystonias

The network of brain regions functionally connected to each lesion location causing dystonia was identified (Figure 2A). Over 80% of lesion locations causing dystonia were functionally connected to a set of common brain regions (Figure 2B, i and ii). Many of these connections were specific to dystonia compared to lesions causing other neurological symptoms (Figure 2B, iii and iv), and the conjunction of sensitive and specific connections was used to derive a ‘general dystonia network’ (Figure 2, v). The major anatomical structures implicated were the basal ganglia, cerebellum, thalamus, claustrum, brainstem (positive connectivity, surviving 4/4 methods), mid/superior temporal gyrus, medial frontal gyrus, and posterior parietal cortex (PPC) (negative connectivity, surviving 3/4 methods). Additional slices and MNI coordinates are presented in Supplementary Files 5 and 6.

### Network connectivity and extent of dystonia in the body

Next, we investigated if greater connectivity to the general dystonia network could predict a greater extent of dystonia in the body (illustrated in Figure 3A). These analyses demonstrated that lesions with stronger connectivity to the general dystonia network had more body parts affected by dystonia (ρ=0.235; p=0.002; Figure 3B) and a wider distribution of dystonia in the body (e.g., generalised dystonia) (F[4,174]=4.37; p=0.002, Figure 3C). These relationships were robust to analytic approach, remaining significant when analysing the number of overlapping voxels, instead of connectivity strength (number of body parts: ρ=0.229; p=0.002; dystonia body distribution: F[4,174]=3.13; p=0.016, Supplementary File 7).

**Figure 3.**
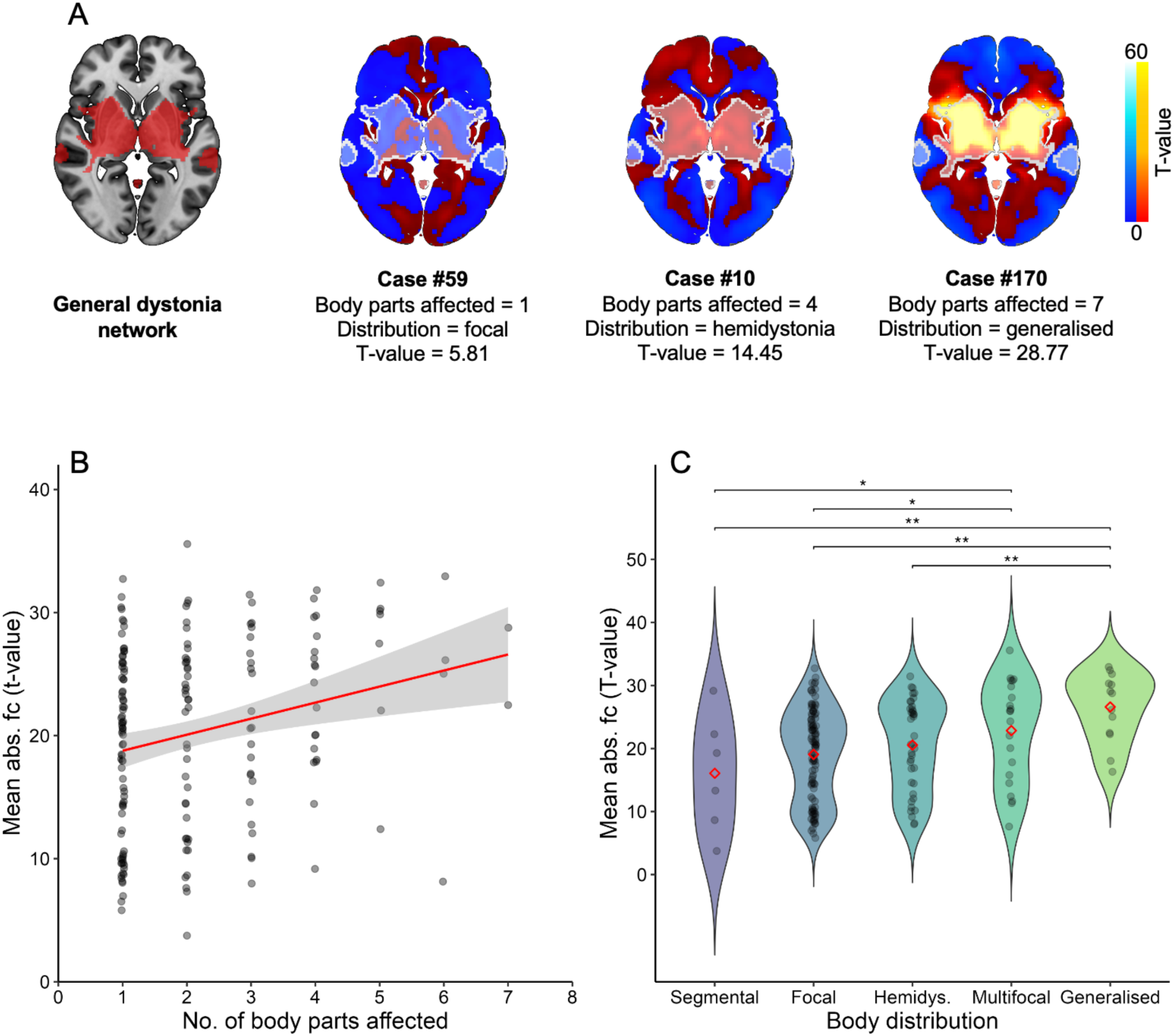
Network connectivity and extent of dystonia in the body. Figure A shows three example cases illustrating the relationship between stronger lesion connectivity (weakest to strongest, left to right) to the general dystonia network (red areas in left brain and outlined in white in three brains on right [cases 59, 10, 170]) and a greater extent of dystonia in the body. The mean absolute functional connectivity t-value of voxels within the general dystonia network is shown. Figures B and C demonstrate this relationship statistically: Figure B shows the significant positive association between a lesion’s connectivity strength to the general dystonia network and the number of body parts affected by dystonia (p=0.002). Figure C shows the relationship between connectivity strength to the network and dystonia body distribution (defined as per prior consensus statements^1,2^) – connectivity strength was significantly higher for cases with generalised or multifocal dystonia, compared to other dystonia body distributions. ** = p<0.01. * = p<0.05.

These results were also specific to connectivity to the dystonia network; Neither lesion size, nor lesion connectivity to brain regions outside the dystonia network, were associated with the number of body parts affected or dystonia body distribution (all p>0.20; see Supplementary File 8 for both analyses).

### Network connectivity and dystonia symptom latency

Unlike the extent of dystonia in the body, the delay between the lesion and the onset of dystonia symptoms was not significantly associated with connectivity strength to the general dystonia network (p = 0.603), or the number of voxels hitting the general dystonia network (p = 0.889). Likewise, there were no voxelwise significant associations between symptom latency and lesion connectivity, either in whole brain analysis or restricted to voxels within the general dystonia network (all voxels p>0.35 FWE-corrected).

### Lesion network mapping by affected body region

We then computed lesion networks for each dystonic body part using the same statistical tests and control groups used to define the general dystonia network. Although lesions across the different dystonia types were strongly connected to the general dystonia network, there was preferential localisation of different hubs of this network, and some body part analyses localised structures outside of the general dystonia network (Figure 4, see Supplementary file 9 for results of the individual tests that were used to define these networks). Certain brain structures, such as the basal ganglia and cerebellum, were present in nearly all dystonia networks. Other structures appeared in some dystonia networks but not others. For example, the sensorimotor cortex was prominent in the cervical dystonia network, while the PPC was consistently localised across the limb dystonia networks (Figure 4).

**Figure 4.**
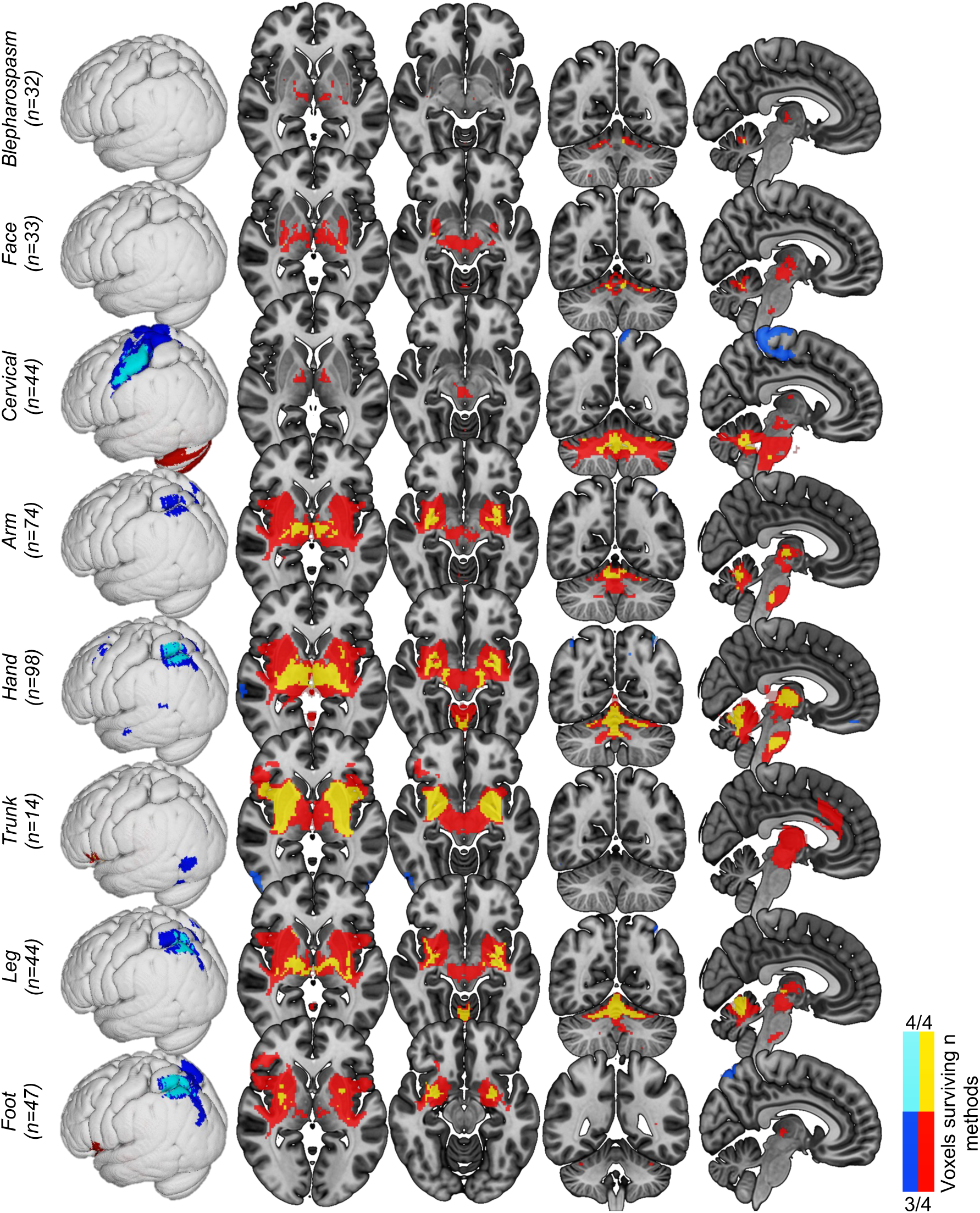
Dystonia networks by affected body parts. The same statistical tests shown in Figure 2, comparing dystonia cases to those from other neurological disorders, were run for each body part affected by dystonia. Voxels surviving ≥3 of these four statistical tests were defined as the lesion network for each body part (red/yellow = positive connectivity; blue/light blue = negative connectivity). T-values are shown for visual purposes, masked to regions p<0.05 FWE corrected. Labels show n of lesions for each group. Note that body parts with a higher n of causal lesions have greater power to demonstrate significant effects.

Next, we statistically compared the lesion networks associated with different body parts and found several significant differences (P_FWE_<0.05). Lesions causing cervical, hand, trunk, and foot dystonia demonstrated connections that were significantly stronger for that dystonia type when compared to other types of dystonia (Figure 5). The dystonia networks by dystonia body distribution (e.g., focal, hemidystonia, etc.)^1,2^ are shown in Supplementary files 9 and 10.

**Figure 5.**
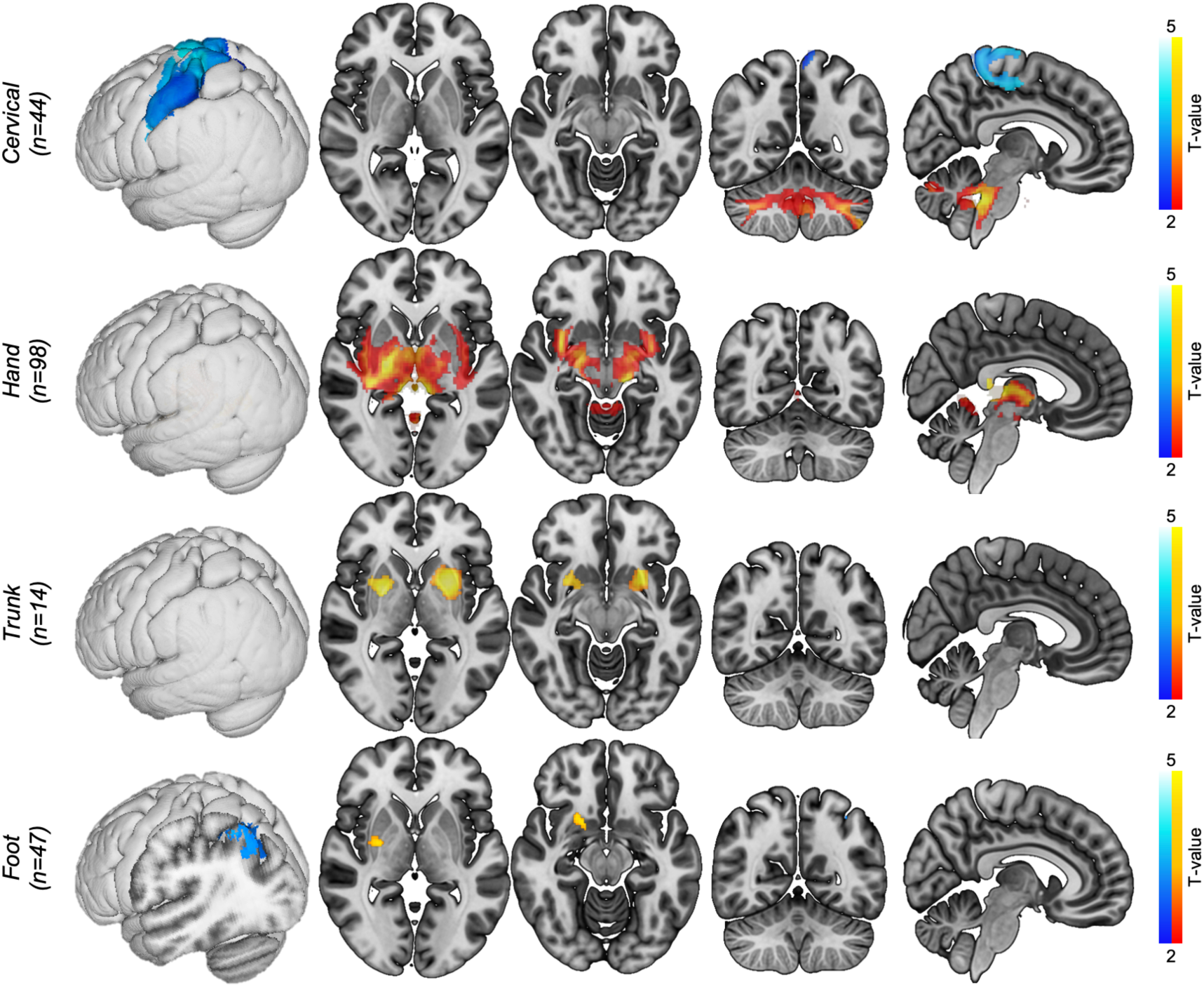
Significant differences between dystonias. ‘Within dystonia’ analyses compared lesion connectivity between cases with dystonia to that body part versus cases without (e.g., 44 lesions causing cervical dystonia versus 135 dystonia lesions not involving cervical dystonia). T-values are shown for visual purposes, masked to regions p<0.05 FWE corrected. Body parts not shown did not have significant findings. Warm colours = positive connectivity; cool colours = negative connectivity.

### Somatotopic representation of dystonic body parts

Given prior evidence implicating the cerebellum in dystonia^24,47,48^ and the established somatotopic organisation of this structure^41,42^, we investigated whether different types of dystonia might map to somatotopically organised cerebellar sub-regions. Specifically, we explored whether the lesion networks from cervical, hand, and foot dystonia (Figure 4) aligned with somatotopic representations of the tongue, hand, and foot in the cerebellum in healthy individuals^41^. There was a clear posterior to anterior organisation for these different dystonia body part networks in the cerebellar lobes (Figure 6A, i, see Supplementary file 11 for additional slices), which resembled the pattern shown previously in Buckner et al.^41^ (Figure 6A, ii). Figure 6B shows the SUIT atlas^44,45^ sub-structures in which these networks fell (See Supplementary file 12 for quantification of voxels within each anatomical sub-structure). Additional analyses demonstrated that these different dystonia body part networks also localised to different sub-regions of the thalamus and putamen (Supplementary file 13).

**Figure 6.**
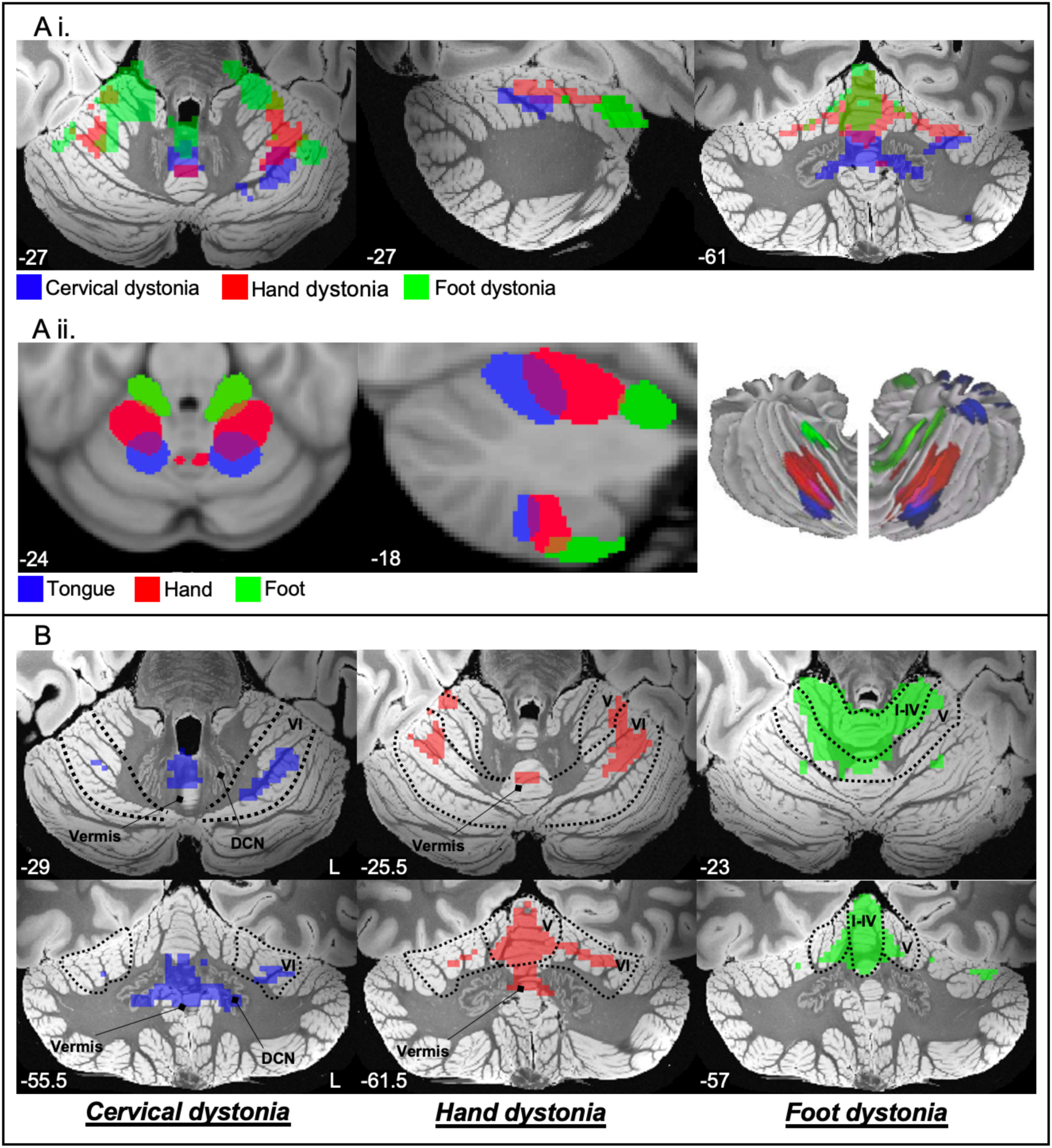
Somatotopic representations of dystonic body parts. Figure A, i. shows that the cervical, hand, and foot dystonia maps resembled a posterior to anterior pattern also demonstrated for the tongue, hand, and foot in healthy participants by Buckner et al., (Figure A, ii). Figure B shows the cerebellar sub-structures implicated between the body part networks. For cervical dystonia, the cluster fell mainly within the vermis, lobule VI, and the deep cerebellar nuclei (DCN). For hand dystonia, the cluster fell mainly within lobule V, lobule VI, and the vermis. For foot dystonia, the cluster fell mainly within lobules I-IV, lobule V, and lobule VI. Figure A. ii. re-produced with permission.

## Discussion

The aim of this study was to leverage causal brain lesions to map the neuroanatomy of dystonia and test whether different types of dystonia mapped to different neural networks. While lesions causing dystonia occurred in heterogeneous locations, they localised to a common cortico-basal ganglia-cerebellar network (‘general dystonia network’). The strength of connectivity between lesion locations and this dystonia network was associated with a greater extent of dystonia in the body. However, the preferential involvement of the hubs of this network varied between the different types of dystonia, which also demonstrated somatotopic organisation in the cerebellum. These findings provide new insight into the brain networks underlying dystonia and its heterogeneous clinical presentation.

### A general brain network of dystonia

Our analysis across all 179 cases of dystonia caused by focal brain lesions localised a brain network primarily encompassing the basal ganglia, cerebellum, thalamus, claustrum, brainstem, and cerebral cortex (Figure 2). This finding corroborates the structures hypothesised by the ‘network model’ of dystonia^7,49^, yet also greatly extends upon this model by localising the network in detail through analysis of a large sample of causal lesions. Notably, this network aligns with the neural pathways connecting the basal ganglia and cerebellum^50,51^. Output pathways of the cerebellum (i.e., deep nuclei) can regulate striatal activity via the thalamus^51^, while output neurons of the basal ganglia project to the cerebellar cortex via the pontine nuclei^50,51^. All of these structures were part of our data-driven lesion-based general dystonia network, consistent with the hypothesis that dystonia arises from dysfunction of this basal ganglia-cerebellar network^50^. Research in animals aligns with this hypothesis, with the basal ganglia and the cerebellum the two primary targets that can induce dystonia-like posturing in rodent models. For example, Calderon et al.^52^ demonstrated that pharmacological manipulation of both structures was required for stress-induced dystonic posturing in a mouse model, and that severing the di-synaptic link between the cerebellum and the basal ganglia through thalamic lesioning alleviated the dystonia. Similarly, sub-clinical lesions of the mouse striatum have been shown to exaggerate cerebellar-induced dystonia posturing, which could in-turn be alleviated by cerebellectomy^53^. These interventions can produce different forms of dystonic posturing in animal models, and the affected pathways may vary between these phenotypes^54^. This agrees with our analyses showing that the major structures of the general dystonia network were implicated to varying degrees across the different types of dystonia, and the suggestion of Jinnah et al.^49^ that in the ‘network model’ of dystonia, the starting point may be different between the dystonia types.

### Brain networks in different manifestations of dystonia

A central mystery of dystonia has been the unexplained variability between patients in the extent of dystonia throughout the body (e.g., focal vs. generalised), and in the body regions in which it occurs. Our results help to explain this mystery in two ways. First, we demonstrated that stronger connectivity to our general dystonia network was associated with more widespread dystonia in the body. This suggests that the greater the disruption to the neural substrates in the brain underpinning dystonia symptoms, the greater number of body parts that are dystonic. This may also provide an account of why dystonia can spread from one part of the body to other parts, with neuroplastic mechanisms impacting additional parts of the network over time. However, this is speculative, and longitudinal studies in dystonia would be needed to confirm this suggestion. Second, our results demonstrate differences in the preferential involvement of the dystonia network hubs between the different dystonia types. This indicates that different hubs are dominant for different dystonia types, and that other hubs can remain relatively unaffected. For example, lesions causing CD were strongly connected to the cerebellum but less so to the basal ganglia, and the opposite pattern was true for lesions causing foot dystonia (Figures 4 and 5). Therefore, for example, CD appears to be a ‘cerebellar dominant’ form of dystonia, while foot dystonia appears to be a ‘basal ganglia dominant’ form of dystonia. This is in agreement with a recent model of the neural structures that may underlie phenotypic differences in dystonia, with the cerebellum proposed to be more involved in cervical and facial dystonias, and the basal ganglia more involved in limb dystonias^54^. Our findings corroborate this suggestion and offer perhaps the clearest evidence yet of the differential role of these two structures in dystonia.

In addition to these subcortical regions in dystonia, our findings also implicated specific cortical regions. Again, their preferential involvement differed between dystonia types, with lesion connectivity to the sensorimotor cortex in CD, but the PPC in limb dystonias. The localisation of the sensorimotor cortex in CD strengthens our earlier finding (with an increased sample size here)^24^ by demonstrating that not only is this region specific compared to other neurological disorders, but also to other types of dystonia (Figure 5), suggesting a unique role of the sensorimotor cortex in CD. Abnormal neural activity in the sensorimotor cortex in CD is well supported by prior evidence^55–57^, and also by the higher proportion of CD patients who show symptom relief through sensory tricks compared with other dystonia types^8^. Conversely, the PPC is involved in spatial orientation and movement of the limbs, forming a part of the ‘reach-to-grasp’ network, and receives strong input from the thalamus^58,59^. The PPC has also been implicated in hand dystonia previously^60,61^, and a recent study demonstrated that a single session of transcranial magnetic stimulation to the somatosensory cortex produced changes in BOLD activity in the PPC that were correlated with writing performance in hand dystonia patients^62^.

### Somatotopic organisation

Our results suggest that cervical, hand, and foot dystonia localise different regions of the cerebellum that align with somatotopic representations^41,42^. Numerous studies have shown that dystonia is associated with disorganisation of motor somatotopy compared to healthy controls, including aberrant activation patterns and variable distances between representations^63–65^. However, to our knowledge, this is the first study to demonstrate distinct somatotopic representations between the different body parts affected by dystonia. This suggests that damage to a particular body part representation in the brain may contribute to the generation of dystonia symptoms in the corresponding body part.

It is also notable that the CD network mapped most strongly to the vermis and deep cerebellar nuclei, whereas the foot dystonia network was more confined to the cerebellar hemispheres (Figure 6). Anatomically, the vermis projects to deep cerebellar nuclei, which then output to the cerebral cortex via the thalamus, and is especially associated with the control of proximal and axial body movements^66,67^. In contrast, the cerebellar hemispheres receive inputs from numerous structures, including projections from the basal ganglia via the pontine nuclei^50,51^. Therefore, these findings align with our above suggestion of CD as cerebellar dominant, and foot dystonia as a basal ganglia dominant form of dystonia.

### Implications for therapy

Our findings appear to align with neural structures and networks that have been shown to mediate therapeutic response in dystonia. First, the globus pallidus interna, the primary treatment target for deep brain stimulation (DBS) for all subtypes of dystonia^68,69^, was a part of our general dystonia network (Figure 2). Two studies have recently shown that therapeutic response to globus pallidus DBS was associated with connectivity to the cerebellum^70,71^, which was also part of our lesion-based general dystonia network. Further, our hand dystonia network strongly implicated the thalamus, and a prior retrospective analysis of 171 task specific focal hand dystonia patients demonstrated significant improvements after ventro-oral thalamotomy, with good or partial response in 97.7% of cases^72^. These and other lesion network mapping data imply a link between causal lesions and the guidance of therapeutic targets^20,73,74^, and our findings provide specific testable targets for new interventions in dystonia. For instance, transcranial magnetic stimulation targeting the somatosensory cortex derived from our earlier lesion network mapping findings^24^ is currently being investigated in the treatment of CD (ACTRN12621000417886^55^). Importantly however, the present findings indicate that some types of dystonia may require different treatment targets. For example, a different cortical region, the PPC, was localised for limb dystonias (Figure 4). These results provide specific and testable hypotheses for neuromodulation in dystonia.

### Limitations

Several limitations should be acknowledged. First, although we conducted a systematic review, there is the possibility of publication bias affecting findings given that lesions in regions previously linked with dystonia may be more likely to be published. Yet as lesion-induced dystonia is relatively rare^75^, collecting a prospective sample is likely not feasible and the current analyses are based on all available data. Second, as we were forced to rely on the diagnosis and reporting of original publication authors, we were not able to clinically confirm the diagnoses. However, each case was reviewed for consistency with lesion-induced dystonia and any incorrect diagnoses would likely add noise into the data, biasing us against the present findings. Third, as in most lesion studies, our cases often involved multiple lesions; therefore, in these cases it is unclear whether dystonia was caused by one lesion or a combination of lesions. It is also difficult to conclusively prove a causal relationship between a lesion and a neurological disorder, especially given the long symptom latency associated with dystonia^8^. Therefore, in some of cases it is possible that the lesion may not have been the cause of dystonia. Yet this is a limitation inherent to retrospective lesion-based analyses, and we only included cases where authors concluded that the lesion was the cause of the dystonia. In addition, again these limitations should theoretically only increase variance and bias us against significant findings. Next, in relation to lesion network mapping, we used 2D lesions rather than 3D lesions from individuals’ MRI scans, but this has been shown to have little impact on the resulting lesion networks^19,76^. Finally, findings are based on functional connectivity data from a normative connectome rather than one matched to the disease under investigation. However, this has been shown to have minimal impact on findings in previous studies^77,78^.

### Conclusions

This study demonstrates a causal link between dystonia and damage to a specific cortico-basal ganglia-cerebellar network. Further, stronger connectivity to this network from causal brain lesions was associated with more widespread dystonia throughout the body, and preferential localisation of the hubs of this network significantly differed between the different dystonia types. These findings indicate that variations in dystonia phenotypes are driven by variations in the neural structures that are affected. The identification of these lesion networks offers specific, testable targets for future neuromodulation trials that can be tailored to the specific dystonia population under investigation.

## Data Availability

All data produced in the present study are available upon reasonable request to the authors

