## Supplementary Files for "Mapping the Neuroanatomy of Dystonia Using Causal Brain Lesions"

**Supplementary file 1. Systematic search syntax**

Search of ‘all fields’ conducted in Pubmed including cases up to July 2022:

(dystonia OR blepharospasm OR meige OR meige’s) AND (lesion OR lesions OR stroke OR infarct* OR ischem* OR ischaem* OR hemorrhag* OR haemorrhag* OR tumor OR tumour OR plaque*).

Search of titles and abstracts and keywords conducted in Embase including cases up to September 2022:

('dystonia':ti,ab,kw OR 'blepharospasm':ti,ab,kw OR 'meige':ti,ab,kw OR 'meige/s':ti,ab,kw) AND ('(lesion':ti,ab,kw OR 'lesions':ti,ab,kw OR 'stroke':ti,ab,kw OR 'infarct*':ti,ab,kw OR 'ischem*':ti,ab,kw OR 'ischaem*':ti,ab,kw OR 'hemorrhag*':ti,ab,kw OR 'haemorrhag*':ti,ab,kw OR 'plaque*':ti,ab,kw OR 'tumor':ti,ab,kw OR 'tumour)':ti,ab,kw).

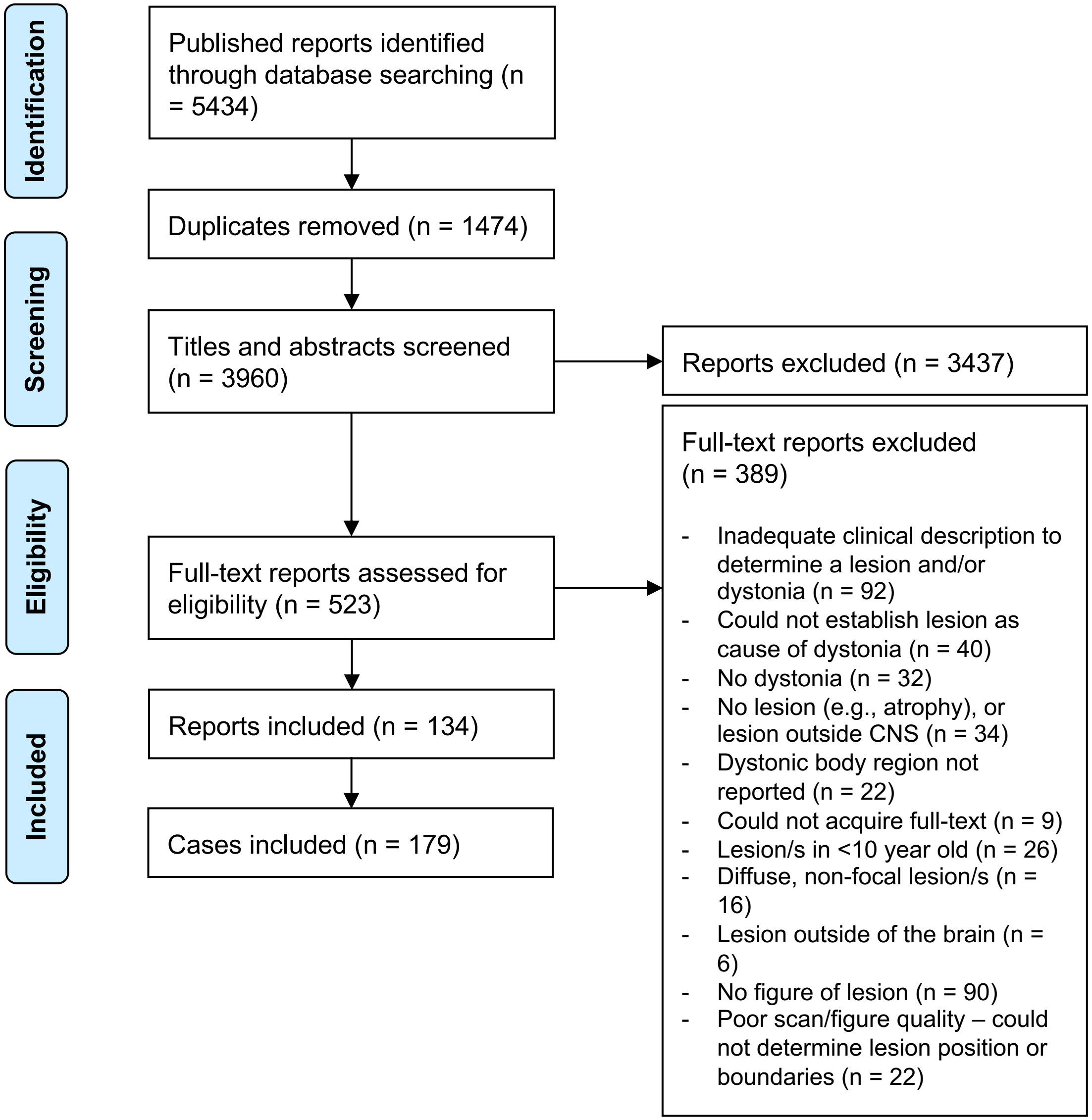

**Supplementary File 2. Systematic search process.** Flowchart adapted from Liberati et al. (2009). There were more cases than published reports because some reports contained multiple cases.

**Supplementary File 3. Post-stroke non-specific neurological disorders dataset description**

500 post-stroke patients were prospectively enrolled from the Turku University Hospital Division of Clinical Neurosciences as part of an on-going project at Turku University Hospital (Turku, Finland). The project was approved by the local ethics committee and hospital district, and conducted according to the principles of the Declaration of Helsinki. All cases were above the age of 18 years, with an acute focal stroke, willing to participate in the study, and gave written informed consent. Cases with diffuse, non-parenchymal or non-focal cerebrovascular events or no visible lesion in clinical brain imaging were excluded. Licensed clinicians specialising in neurology performed clinical evaluations in consultation with a consultant neurologist (J.J.). Cases without dystonia were included here as a post-stroke comparison group (n = 499).

**Supplementary file 4.** Clinical characteristics of the included cases.

| Case | Study | StudyCase | Age range | Sex | BodyDistribution | Bleph. | Face | Cervical | Arm | Hand | Trunk | Leg | Foot | Variability |
|---|---|---|---|---|---|---|---|---|---|---|---|---|---|---|
| 1 | Aasfara (2012) | | 15-19 | Male | Focal | No | No | No | Yes | Yes | No | No | No | Persistent |
| 2 | Aguirregomozcorta (2008) | | 35-39 | Female | Hemidystonia | No | No | No | Yes | Yes | No | Yes | No | Paroxysmal |
| 3 | Akin (2014) | | 65-69 | Female | Focal | No | Yes | No | No | No | No | No | No | Persistent |
| 4 | Alarcon (2001) | 1 | 40-44 | Female | Focal | No | No | No | Yes | Yes | No | No | No | Persistent |
| 5 | Alvarez (2014) | 1 | 35-39 | Male | Focal | No | No | No | Yes | Yes | No | No | No | Persistent |
| 6 | Apaydin (1998) | 1 | 70-74 | Female | Focal | No | No | No | Yes | Yes | No | No | No | Persistent |
| 7 | Apaydin (1998) | 2 | 60-64 | Male | Hemidystonia | No | No | No | Yes | Yes | No | Yes | No | Persistent |
| 8 | Aramideh (1996) | | 45-49 | Female | Focal | Yes | No | No | No | No | No | No | No | Persistent |
| 9 | Azevedo (2001) | | 25-29 | Female | Focal | No | No | No | Yes | Yes | No | No | No | Persistent |
| 10 | Baldacci (2010) | | 45-49 | Male | Hemidystonia | No | No | No | Yes | Yes | No | Yes | Yes | Paroxysmal |
| 11 | Batla (2015) | 1 | 55-59 | Female | Focal | No | No | Yes | No | No | No | No | No | NR |
| 12 | Batla (2015) | 2 | 30-34 | Male | Focal | No | No | Yes | No | No | No | No | No | NR |
| 13 | Batla (2015) | 5 | 55-59 | Male | Multifocal | No | No | Yes | Yes | No | No | No | No | NR |
| 14 | Batla (2015) | 7 | 25-29 | Male | Focal | No | No | Yes | No | No | No | No | No | NR |
| 15 | Berkovic (1987) | 3 | 45-49 | Male | Generalized | No | Yes | Yes | No | No | Yes | Yes | Yes | Persistent |
| 16 | Bogdanova-Mihaylova (2017) | | 40-44 | Female | Focal | No | No | No | Yes | Yes | No | No | No | Paroxysmal |
| 17 | Braga-Neto (2008) | 1 | 25-29 | Female | Hemidystonia | No | No | No | Yes | Yes | No | No | No | Persistent |
| 18 | Brett (1981) | | 10-14 | Female | Hemidystonia | No | No | No | Yes | Yes | No | No | Yes | Persistent |
| 19 | Buerger (2014) | | 20-24 | Male | Hemidystonia | No | Yes | No | Yes | No | No | No | No | Paroxysmal |
| 20 | Burguera (2001) | 1 | 45-49 | Male | Focal | No | No | No | No | Yes | No | No | No | Action_specific |
| 21 | Camac (1990) | | 50-54 | Female | Hemidystonia | No | Yes | No | Yes | Yes | No | Yes | No | Paroxysmal |
| 22 | Chang (2002) | 1 | 20-24 | Male | Focal | No | No | Yes | No | No | No | No | No | Persistent |
| 23 | Chauhan (2009) | 1 | 45-49 | Female | Focal | Yes | No | No | No | No | No | No | No | Persistent |
| 24 | Cho (2000) | 1 | 65-69 | Female | Focal | No | No | No | No | Yes | No | No | No | Persistent |
| 25 | Choi (1993) | 1 | 55-59 | Male | Focal | No | No | No | No | Yes | No | No | No | Persistent |
| 26 | Choi (1993) | 3 | 15-19 | Male | Focal | No | No | No | No | No | No | No | Yes | Persistent |
| 27 | Choi (2015) | | 65-69 | Male | Multifocal | No | Yes | No | Yes | Yes | No | Yes | Yes | Persistent |
| 28 | Comoglu (2002) | 1 | 60-64 | Male | Focal | Yes | No | No | No | No | No | No | No | Persistent |
| 29 | Day (1986) | | 75-79 | Female | Segmental | Yes | Yes | No | No | No | No | No | No | Persistent |
| 30 | Defebvre (1995) | 1 | 60-64 | Female | Multifocal | No | No | No | Yes | Yes | No | Yes | Yes | Persistent |
| 31 | Deleu (2000) | | 15-19 | Female | Focal | No | No | No | No | Yes | No | No | No | Persistent |
| 32 | Demierre (1983) | 3 | 15-19 | Male | Hemidystonia | No | No | No | Yes | Yes | No | Yes | Yes | Persistent |

| | | | | | | | | | | | | | |
|---|---|---|---|---|---|---|---|---|---|---|---|---|---|
| 33 | Di Capua (2001) | | 10-14 | Female | Focal | No | No | No | No | Yes | No | No | No | Action_specific |
| 34 | Dietrichs (2000) | | 70-74 | Female | Focal | No | Yes | No | No | No | No | No | No | Persistent |
| 35 | Dinkelbach (2015) | | 45-49 | Female | Multifocal | No | No | Yes | No | No | Yes | No | No | Persistent |
| 36 | Esteban Munoz (1996) | | 45-49 | Female | Focal | No | No | No | Yes | Yes | No | No | No | Persistent |
| 37 | Franzini (2009) | | 30-34 | Female | Focal | No | No | No | No | No | No | No | Yes | Persistent |
| 38 | Fuller (2013) | | 40-44 | Male | Generalized | No | No | Yes | Yes | Yes | Yes | Yes | No | Persistent |
| 39 | Galvez-Ruiz (2014) | 1 | | | Focal | Yes | No | No | No | No | No | No | No | NR |
| 40 | Ghika (1994) | 1 | 90-94 | Female | Focal | No | No | No | No | Yes | No | No | No | Persistent |
| 41 | Ghika (1994) | 2 | 60-64 | Male | Focal | No | No | No | No | Yes | No | No | No | Persistent |
| 42 | Ghika (1994) | 3 | 30-34 | Male | Focal | No | No | No | Yes | Yes | No | No | No | Persistent |
| 43 | Gilbert (2012) | | 30-34 | Female | Focal | Yes | No | No | No | No | No | No | No | Persistent |
| 44 | Gille (1996) | | 65-69 | Male | Hemidystonia | No | No | No | Yes | Yes | No | Yes | Yes | Persistent |
| 45 | Grandas (1989) | | 35-39 | Male | Generalized | No | No | No | Yes | Yes | Yes | Yes | Yes | Persistent |
| 46 | Grandas (2004) | | 65-69 | Female | Focal | Yes | No | No | No | No | No | No | No | Persistent |
| 47 | Guak (2002) | | 20-24 | Female | Multifocal | No | Yes | No | Yes | No | No | No | No | Paroxysmal |
| 48 | Hamasaki (2008) | 1 | 55-59 | Male | Hemidystonia | No | No | No | Yes | Yes | No | Yes | No | Persistent |
| 49 | Hamasaki (2008) | 2 | 70-74 | Female | Hemidystonia | Yes | Yes | No | Yes | No | No | Yes | No | Persistent |
| 50 | Hawker (1990) | 1 | 55-59 | Female | Multifocal | No | No | No | Yes | Yes | No | Yes | Yes | Persistent |
| 51 | Hawker (1990) | 3 | 45-49 | Male | Focal | No | No | No | No | No | No | No | Yes | Persistent |
| 52 | Hsieh (2014) | | 70-74 | Male | Focal | Yes | No | No | No | No | No | No | No | Persistent |
| 53 | Isaac (1989) | | 25-29 | Male | Focal | No | No | Yes | No | No | No | No | No | Persistent |
| 54 | Jacob (1995) | | 40-44 | Male | Segmental | Yes | Yes | No | No | No | No | No | No | Persistent |
| 55 | Jankovic (1983) | 1 | 55-59 | Female | Focal | Yes | No | No | No | No | No | No | No | Persistent |
| 56 | Jankovic (1983) | 4 | 50-54 | Female | Segmental | Yes | Yes | No | No | No | No | No | No | Persistent |
| 57 | Jankovic (1986) | | 50-54 | Male | Focal | Yes | No | No | No | No | No | No | No | Persistent |
| 58 | Jung (2016) | | 45-49 | Female | Hemidystonia | No | No | No | No | No | No | Yes | Yes | Paroxysmal |
| 59 | Kajimoto (2004) | 1 | 80-84 | Female | Focal | No | No | Yes | No | No | No | No | No | Persistent |
| 60 | Karsidag (1998) | 1 | 65-69 | Female | Focal | No | No | No | No | Yes | No | No | No | Action_specific |
| 61 | Karsidag (1998) | 2 | 60-64 | Female | Focal | No | No | No | No | Yes | No | No | No | Persistent |
| 62 | Karsidag (1998) | 4 | 45-49 | Male | Focal | No | No | No | No | Yes | No | No | No | Persistent |
| 63 | Karsidag (1998) | 5 | 60-64 | Male | Focal | No | No | No | No | Yes | No | No | No | Persistent |
| 64 | Karsidag (1998) | 7 | 50-54 | Female | Generalized | No | No | Yes | No | Yes | Yes | Yes | No | Persistent |
| 65 | Karsidag (1998) | 8 | 65-69 | Female | Focal | No | No | No | No | Yes | No | No | No | Persistent |
| 66 | Karsidag (1998) | 9 | 50-54 | Female | Focal | No | No | No | No | Yes | No | No | No | Persistent |

| | | | | | | | | | | | | | | |
|---|---|---|---|---|---|---|---|---|---|---|---|---|---|---|
| 67 | Keane (1985) | | 30-34 | Male | Multifocal | Yes | No | No | No | Yes | No | No | No | Persistent |
| 68 | Kemp (2012) | | 40-44 | Male | Focal | No | No | No | No | No | No | Yes | No | Action_specific |
| 69 | Kim (2009) | | 70-74 | Male | Focal | No | Yes | No | No | No | No | No | No | Persistent |
| 70 | Kirton (2001) | | 60-64 | Female | Segmental | Yes | Yes | Yes | No | No | No | No | No | Paroxysmal |
| 71 | Koc (2010) | | 10-14 | Male | Hemidystonia | No | No | No | Yes | No | No | No | No | Persistent |
| 72 | Koch (2006) | | 60-64 | Male | Focal | No | No | No | No | No | No | Yes | Yes | Paroxysmal |
| 73 | Krauss, Mohadjer, Wakhloo (1991) | | 35-39 | Female | Multifocal | Yes | Yes | Yes | No | No | No | Yes | No | Persistent |
| 74 | Krystkowiak (1998) | 5 | 55-59 | | Focal | No | No | No | Yes | Yes | No | No | No | NR |
| 75 | Kulisevsky (1993) | | 55-59 | Female | Focal | Yes | No | No | No | No | No | No | No | Paroxysmal |
| 76 | Kuoppamaki (2002) | 2 | 20-24 | Female | Generalized | No | Yes | No | Yes | Yes | Yes | Yes | Yes | Persistent |
| 77 | Lambrecq (2010) | | 20-24 | Male | Multifocal | Yes | No | Yes | No | No | No | No | No | Persistent |
| 78 | Larumbe (1993) | | 10-14 | Female | Generalized | Yes | No | No | Yes | No | Yes | Yes | No | Action_specific |
| 79 | Lauterbach (1994) | | 60-64 | Female | Focal | Yes | No | No | No | No | No | No | No | Persistent |
| 80 | LeDoux (2003) | 1 | 55-59 | Male | Focal | No | No | Yes | No | No | No | No | No | Persistent |
| 81 | LeDoux (2003) | 3 | 65-69 | Female | Focal | No | No | Yes | No | No | No | No | No | Persistent |
| 82 | LeDoux (2003) | 4 | 70-74 | Male | Focal | No | No | Yes | No | No | No | No | No | Persistent |
| 83 | Lee, Lee (1996) | | 30-34 | Male | Multifocal | No | Yes | No | No | Yes | No | No | No | Persistent |
| 84 | Lee, Rinne (1994) | 1 | 10-14 | Female | Hemidystonia | No | No | No | Yes | Yes | No | Yes | No | Persistent |
| 85 | Leenders (1986) | | 40-44 | Male | Multifocal | Yes | No | No | Yes | Yes | No | Yes | No | Persistent |
| 86 | Lehericy (1996) | 3 | 55-59 | Female | Multifocal | No | No | No | No | Yes | No | Yes | Yes | Persistent |
| 87 | Lehericy (1996) | 5 | 70-74 | Male | Focal | No | No | No | Yes | Yes | No | No | No | Persistent |
| 88 | Lehericy (2001) | 1 | 60-64 | | Focal | No | No | No | No | Yes | No | No | No | Persistent |
| 89 | Lehericy (2001) | 5 | 45-49 | | Focal | No | No | No | No | Yes | No | No | No | Persistent |
| 90 | Lehericy (2001) | 6 | 65-69 | | Focal | No | No | No | No | Yes | No | No | No | Persistent |
| 91 | Lehericy (2001) | 7 | 50-54 | | Focal | No | No | No | No | Yes | No | No | No | Persistent |
| 92 | Lera (2000) | 2 | 75-79 | Female | Focal | No | No | No | Yes | Yes | No | No | No | Persistent |
| 93 | Lera (2000) | 7 | 70-74 | Female | Focal | No | No | No | Yes | No | No | No | No | Persistent |
| 94 | Liepert (2003) | 1 | 55-59 | Male | Focal | No | No | No | No | Yes | No | No | No | Action_specific |
| 95 | Loher (2009) | 1 | 30-34 | Male | Multifocal | No | Yes | Yes | Yes | Yes | No | Yes | No | Persistent |
| 96 | Loher (2009) | 2 | 40-44 | Male | Hemidystonia | No | No | Yes | Yes | No | No | Yes | No | Persistent |
| 97 | Loher (2009) | 4 | 55-59 | Male | Hemidystonia | Yes | No | Yes | Yes | No | No | No | No | Persistent |
| 98 | Mahant (2003) | | 35-39 | Female | Focal | No | No | No | No | No | No | Yes | Yes | Persistent |
| 99 | Marsden (1985) | 8 | 10-14 | Male | Hemidystonia | No | No | No | Yes | Yes | No | Yes | No | Persistent |
| 100 | Marsden (1985) | 10 | 75-79 | Female | Focal | No | No | No | No | Yes | No | No | No | Persistent |

| | | | | | | | | | | | | | | |
|---|---|---|---|---|---|---|---|---|---|---|---|---|---|---|
| 101 | Marsden (1985) | 11 | 20-24 | Female | Hemidystonia | No | No | No | Yes | Yes | No | No | Yes | Persistent |
| 102 | Marsden (1985) | 22 | 60-64 | Female | Hemidystonia | No | No | No | Yes | Yes | No | Yes | No | Persistent |
| 103 | Micheli (1998) | | 10-14 | Female | Hemidystonia | No | No | No | Yes | Yes | No | No | Yes | Persistent |
| 104 | Miletic (2015) | | 70-74 | Female | Hemidystonia | No | No | No | Yes | No | No | Yes | Yes | Persistent |
| 105 | Miranda (1998) | | 50-54 | Male | Focal | Yes | No | No | No | No | No | No | No | Persistent |
| 106 | Molho (1993) | 1 | 65-69 | Female | Focal | No | No | Yes | No | No | No | No | No | Persistent |
| 107 | Molho (1993) | 2 | 40-44 | Female | Focal | No | No | Yes | No | No | No | No | No | Persistent |
| 108 | Munchau (2000) | 1 | 25-29 | Male | Hemidystonia | No | No | No | Yes | Yes | No | Yes | Yes | Persistent |
| 109 | Munchau (2000) | 3 | 25-29 | Male | Hemidystonia | No | No | Yes | Yes | Yes | No | No | Yes | Persistent |
| 110 | Munchau (2000) | 4 | 25-29 | Male | Focal | No | No | No | No | No | No | Yes | Yes | Persistent |
| 111 | Nishimura (2014) | | 60-64 | Male | Hemidystonia | No | No | No | Yes | Yes | No | No | Yes | Persistent |
| 112 | Nociti (2011) | 1 | 30-34 | Female | Focal | Yes | No | No | No | No | No | No | No | Persistent |
| 113 | Nociti (2011) | 2 | 45-49 | Male | Focal | Yes | No | No | No | No | No | No | No | Persistent |
| 114 | O'Rourke (2006) | | 35-39 | Female | Multifocal | Yes | No | Yes | No | No | No | No | No | Paroxysmal |
| 115 | Olgiati (2016) | 1 | 15-19 | Male | Generalized | No | No | Yes | Yes | Yes | Yes | Yes | Yes | Persistent |
| 116 | Panda (2014) | | 10-14 | Female | Focal | No | No | No | No | Yes | No | No | No | Persistent |
| 117 | Pandey (2014) | | 15-19 | Female | Hemidystonia | No | No | No | No | Yes | No | No | Yes | Persistent |
| 118 | Persing (1990) | | 55-59 | Female | Segmental | Yes | Yes | No | No | No | No | No | No | Persistent |
| 119 | Pettigrew (1985) | 16 | 60-64 | Male | Hemidystonia | No | No | No | No | Yes | No | No | Yes | NR |
| 120 | Plant (1989) | | 30-34 | Female | Focal | No | No | Yes | No | No | No | No | No | Persistent |
| 121 | Powers (1985) | | 60-64 | Male | Multifocal | Yes | No | No | No | Yes | No | No | No | Persistent |
| 122 | Rana (2014) | | 65-69 | Female | Hemidystonia | No | No | No | Yes | Yes | No | No | Yes | Persistent |
| 123 | Riley (1996) | | 65-69 | Male | Focal | No | Yes | No | No | No | No | No | No | Paroxysmal |
| 124 | Sabrie (2015) | | 25-29 | Male | Focal | No | No | No | Yes | Yes | No | No | No | Paroxysmal |
| 125 | Salerno (1993) | | 40-44 | Female | Generalized | No | Yes | Yes | Yes | No | Yes | No | No | Persistent |
| 126 | Schulze-Bonhage (1995) | | 40-44 | Male | Focal | No | No | Yes | No | No | No | No | No | Persistent |
| 127 | Schwartz (1995) | | 60-64 | Male | Focal | No | Yes | Yes | No | No | No | No | No | Persistent |
| 128 | Seet (2005) | | 65-69 | Male | Segmental | No | No | Yes | Yes | No | No | No | No | Paroxysmal |
| 129 | Shen (2016) | | 20-24 | Female | Focal | No | No | No | No | Yes | No | No | No | Persistent |
| 130 | Singer (1998) | | 65-69 | Female | Focal | Yes | No | No | No | No | No | No | No | Persistent |
| 131 | Takahashi (2009) | | 40-44 | Female | Focal | No | No | No | No | Yes | No | No | No | Persistent |
| 132 | Tamburin (2005) | 2 | 35-39 | Female | Multifocal | No | No | No | Yes | Yes | No | Yes | Yes | Paroxysmal |
| 133 | Tan (2005) | | 40-44 | Male | Hemidystonia | No | No | No | Yes | Yes | No | Yes | Yes | Diurnal |
| 134 | Tranchant (1991) | | 50-54 | Female | Focal | No | No | Yes | No | No | No | No | No | NR |

| | | | | | | | | | | | | | | |
|---|---|---|---|---|---|---|---|---|---|---|---|---|---|---|
| 135 | Trankle (1997) | | 25-29 | Male | Focal | No | No | No | No | Yes | No | No | No | NR |
| 136 | Trompetto (2012) | 1 | 50-54 | | Hemidystonia | No | No | No | Yes | No | No | No | Yes | Persistent |
| 137 | Trompetto (2012) | 3 | 60-64 | | Hemidystonia | No | No | No | Yes | No | No | No | Yes | Persistent |
| 138 | Trompetto (2012) | 6 | 45-49 | | Hemidystonia | No | No | No | Yes | Yes | No | No | Yes | Persistent |
| 139 | Trompetto (2012) | 7 | 70-74 | | Focal | No | No | No | No | Yes | No | No | No | Persistent |
| 140 | Usmani (2011) | 1 | 35-39 | Male | Focal | No | No | Yes | No | No | No | No | No | Paroxysmal |
| 141 | Verghese (1999) | | 60-64 | Female | Focal | Yes | No | No | No | No | No | No | No | Action_specific |
| 142 | Vidailhet (1999) | 1 | 50-54 | Male | Hemidystonia | No | Yes | No | Yes | Yes | No | No | No | Persistent |
| 143 | Vidailhet (1999) | 3 | 50-54 | Female | Focal | No | No | No | No | Yes | No | No | No | Persistent |
| 144 | Vidailhet (1999) | 4 | 55-59 | Male | Hemidystonia | No | No | No | Yes | Yes | No | No | Yes | Action_specific |
| 145 | Vidailhet (1999) | 5 | 60-64 | Male | Focal | No | No | No | No | Yes | No | No | No | Persistent |
| 146 | Vidailhet (1999) | 7 | 55-59 | Male | Focal | No | No | No | No | Yes | No | No | No | Persistent |
| 147 | Wali (2001) | | 25-29 | Female | Focal | Yes | No | No | No | No | No | No | No | Persistent |
| 148 | Walker (2015) | | 55-59 | Male | Generalized | No | No | Yes | Yes | No | Yes | No | No | Persistent |
| 149 | Wolz (2008) | | 40-44 | Male | Generalized | No | Yes | Yes | No | No | Yes | No | No | Persistent |
| 150 | Wu (1992) | | 45-49 | Female | Multifocal | No | No | Yes | Yes | Yes | No | No | Yes | Persistent |
| 151 | Zadro (2008) | 1 | 45-49 | Female | Focal | No | No | Yes | No | No | No | No | No | Persistent |
| 152 | Garbin Di Luca (2019) | | 50-54 | Male | Multifocal | No | No | Yes | Yes | Yes | No | Yes | Yes | Paroxysmal |
| 153 | Pandey (2018) | 7 | 55-59 | Male | Focal | No | Yes | No | No | No | No | No | No | Persistent |
| 154 | Pandey (2018) | 9 | 60-64 | Male | Focal | No | Yes | No | No | No | No | No | No | Persistent |
| 155 | Pandey (2018) | 11 | 65-69 | Female | Focal | No | Yes | No | No | No | No | No | No | Persistent |
| 156 | Shin (2019) | | 55-59 | Female | Focal | No | No | No | No | No | No | Yes | Yes | Persistent |
| 157 | Szejko (2020) | | 35-39 | Male | Multifocal | No | No | No | No | Yes | No | No | Yes | Persistent |
| 158 | Hamed (2020) | 6 | 60-64 | Female | Focal | No | Yes | No | No | No | No | No | No | Persistent |
| 159 | Li (2019) | 2 | 15-19 | Male | Hemidystonia | No | No | No | Yes | Yes | No | No | Yes | Persistent |
| 160 | Li (2019) | 3 | 25-29 | Female | Generalized | No | Yes | Yes | Yes | Yes | Yes | Yes | Yes | Persistent |
| 161 | Li (2019) | 5 | 35-39 | Male | Focal | No | No | No | No | Yes | No | No | No | Persistent |
| 162 | Sul (2019) | | 40-44 | Male | Focal | No | No | No | No | Yes | No | No | No | Persistent |
| 163 | Ghosh (2020) | | 25-29 | Female | Focal | No | No | No | Yes | Yes | No | No | No | Persistent |
| 164 | Guimarães Rocha (2021) | 1 | 50-54 | Female | Hemidystonia | No | Yes | Yes | Yes | Yes | No | Yes | Yes | Persistent |
| 165 | Guimarães Rocha (2021) | 2 | 10-14 | Female | Hemidystonia | No | Yes | No | Yes | Yes | No | Yes | Yes | Persistent |
| 166 | Han (2020) | | 40-44 | Male | Multifocal | No | Yes | No | No | Yes | No | No | No | Persistent |
| 167 | Meloni (2021) | | 40-44 | Male | Hemidystonia | No | No | No | Yes | Yes | No | Yes | Yes | Persistent |
| 168 | Owen (2022) | | 35-39 | | Generalized | Yes | No | Yes | Yes | Yes | Yes | Yes | No | Persistent |

| | | | | | | | | | | | | | | |
|---|---|---|---|---|---|---|---|---|---|---|---|---|---|---|
| 169 | Umeh (2012) | | 65-69 | Male | Hemidystonia | No | No | No | Yes | Yes | No | No | Yes | Persistent |
| 170 | Wu, Zheng (2020) | | 15-19 | Female | Generalized | No | Yes | Yes | Yes | Yes | Yes | Yes | Yes | NR |
| 171 | Wu, Su (2020) | | 15-19 | Female | Focal | No | No | No | Yes | Yes | No | No | No | Persistent |
| 172 | Xu (2020) | | 70-74 | Male | Focal | No | No | No | No | Yes | No | No | No | NR |
| 173 | Yu (2021) | | 20-24 | Female | Multifocal | No | No | No | Yes | Yes | No | Yes | Yes | Persistent |
| 174 | Aldosary (2021) | 1 | 10-14 | Male | Focal | No | No | No | Yes | Yes | No | No | No | Action_specific |
| 175 | Smith (2021) | | 35-39 | Male | Focal | No | Yes | No | No | No | No | No | No | Persistent |
| 176 | Funabe (2014) | | 60-64 | Male | Focal | No | No | Yes | No | No | No | No | No | Persistent |
| 177 | Kim, Lee (2007) | | 35-39 | Male | Focal | No | No | Yes | No | No | No | No | No | Persistent |
| 178 | Walker (2007) | | 40-44 | Male | Focal | No | No | Yes | No | No | No | No | No | Persistent |
| 179 | Trompetto (2012) | 9 | 80-84 | | Hemidystonia | No | No | No | Yes | Yes | No | No | Yes | Persistent |

| Case | Study | LatencyWeeks | DystonicTremor | AssociatedTremor | SensoryTrick |
|---|---|---|---|---|---|
| 1 | Aasfara (2012) | 2 | No | No | No |
| 2 | Aguirregomozcorta (2008) | | No | No | No |
| 3 | Akin (2014) | .01 | No | No | Yes |
| 4 | Alarcon (2001) | .5 | Yes | No | |
| 5 | Alvarez (2014) | 17 | Yes | Yes | |
| 6 | Apaydin (1998) | 13 | Yes | No | |
| 7 | Apaydin (1998) | 9 | No | No | |
| 8 | Aramideh (1996) | | No | No | |
| 9 | Azevedo (2001) | | No | No | |
| 10 | Baldacci (2010) | | No | No | No |
| 11 | Batla (2015) | | Yes | No | Yes |
| 12 | Batla (2015) | | No | No | |
| 13 | Batla (2015) | | Yes | No | |
| 14 | Batla (2015) | | Yes | No | |
| 15 | Berkovic (1987) | | No | No | No |
| 16 | Bogdanova-Mihaylova (2017) | 13 | No | No | No |
| 17 | Braga-Neto (2008) | | No | No | |
| 18 | Brett (1981) | 1 | No | No | |
| 19 | Buerger (2014) | 3 | No | No | |
| 20 | Burguera (2001) | 1 | No | No | |
| 21 | Camac (1990) | | No | No | Yes |
| 22 | Chang (2002) | 156 | No | No | Yes |
| 23 | Chauhan (2009) | | No | No | |
| 24 | Cho (2000) | .01 | Yes | No | |
| 25 | Choi (1993) | 52 | No | No | |
| 26 | Choi (1993) | 4 | No | No | |
| 27 | Choi (2015) | | No | No | |
| 28 | Comoglu (2002) | .01 | No | No | |
| 29 | Day (1986) | | No | Yes | |
| 30 | Defebvre (1995) | 3 | No | No | |
| 31 | Deleu (2000) | | No | No | No |
| 32 | Demierre (1983) | 4 | No | No | |

| | | | | | |
|---|---|---|---|---|---|
| 33 | Di Capua (2001) | | No | No | No |
| 34 | Dietrichs (2000) | | Yes | No | Yes |
| 35 | Dinkelbach (2015) | | No | No | Yes |
| 36 | Esteban Munoz (1996) | .01 | Yes | No | |
| 37 | Franzini (2009) | | No | No | No |
| 38 | Fuller (2013) | | No | No | No |
| 39 | Galvez-Ruiz (2014) | | NR | NR | |
| 40 | Ghika (1994) | | Yes | Yes | Yes |
| 41 | Ghika (1994) | | No | No | |
| 42 | Ghika (1994) | | Yes | Yes | |
| 43 | Gilbert (2012) | | No | No | |
| 44 | Gille (1996) | 34 | No | No | |
| 45 | Grandas (1989) | | Yes | No | No |
| 46 | Grandas (2004) | .01 | No | No | |
| 47 | Guak (2002) | | No | No | |
| 48 | Hamasaki (2008) | 13 | No | No | |
| 49 | Hamasaki (2008) | 14 | No | No | |
| 50 | Hawker (1990) | | No | No | |
| 51 | Hawker (1990) | | Yes | Yes | |
| 52 | Hsieh (2014) | | No | No | |
| 53 | Isaac (1989) | 208 | No | No | No |
| 54 | Jacob (1995) | | No | No | |
| 55 | Jankovic (1983) | 52 | No | No | |
| 56 | Jankovic (1983) | 156 | No | No | |
| 57 | Jankovic (1986) | 1.5 | No | No | |
| 58 | Jung (2016) | 21 | No | No | No |
| 59 | Kajimoto (2004) | 1.5 | No | No | |
| 60 | Karsidag (1998) | 39 | No | No | |
| 61 | Karsidag (1998) | 104 | No | No | |
| 62 | Karsidag (1998) | 156 | Yes | No | |
| 63 | Karsidag (1998) | 130 | No | No | |
| 64 | Karsidag (1998) | 4 | No | No | |
| 65 | Karsidag (1998) | 43 | No | No | |
| 66 | Karsidag (1998) | 4 | Yes | No | |

| | | | | | |
|---|---|---|---|---|---|
| 67 | Keane (1985) | 26 | Yes | No | |
| 68 | Kemp (2012) | 39 | No | No | |
| 69 | Kim (2009) | | No | No | |
| 70 | Kirton (2001) | 104 | No | No | |
| 71 | Koc (2010) | 156 | Yes | Yes | No |
| 72 | Koch (2006) | | No | No | No |
| 73 | Krauss, Mohadjer, Wakhloo (1991) | 5 | No | No | |
| 74 | Krystkowiak (1998) | 30 | No | No | |
| 75 | Kulisevsky (1993) | 13 | No | No | |
| 76 | Kuoppamaki (2002) | 8 | No | No | |
| 77 | Lambrecq (2010) | | No | No | |
| 78 | Larumbe (1993) | .5 | No | No | |
| 79 | Lauterbach (1994) | .01 | No | No | |
| 80 | LeDoux (2003) | .1 | No | No | |
| 81 | LeDoux (2003) | .5 | No | Yes | |
| 82 | LeDoux (2003) | .1 | No | No | |
| 83 | Lee, Lee (1996) | 4 | Yes | No | |
| 84 | Lee, Rinne (1994) | 52 | No | No | |
| 85 | Leenders (1986) | | No | No | |
| 86 | Lehericy (1996) | 3 | No | No | |
| 87 | Lehericy (1996) | 4 | No | No | |
| 88 | Lehericy (2001) | | No | No | |
| 89 | Lehericy (2001) | | No | No | |
| 90 | Lehericy (2001) | | No | No | |
| 91 | Lehericy (2001) | | Yes | No | |
| 92 | Lera (2000) | 52 | No | No | |
| 93 | Lera (2000) | .5 | No | No | |
| 94 | Liepert (2003) | | No | No | |
| 95 | Loher (2009) | 13 | Yes | No | |
| 96 | Loher (2009) | 17 | No | No | |
| 97 | Loher (2009) | 4 | Yes | Yes | |
| 98 | Mahant (2003) | | No | No | No |
| 99 | Marsden (1985) | 104 | No | No | |
| 100 | Marsden (1985) | 8 | No | No | |

| | | | | | |
|---|---|---|---|---|---|
| 101 | Marsden (1985) | 8 | Yes | No | |
| 102 | Marsden (1985) | 156 | No | No | |
| 103 | Micheli (1998) | 3 | Yes | No | No |
| 104 | Miletic (2015) | .1 | No | No | |
| 105 | Miranda (1998) | 56 | No | No | |
| 106 | Molho (1993) | | Yes | No | Yes |
| 107 | Molho (1993) | | No | No | Yes |
| 108 | Munchau (2000) | 156 | No | No | |
| 109 | Munchau (2000) | | No | No | |
| 110 | Munchau (2000) | | No | No | |
| 111 | Nishimura (2014) | 1 | No | No | |
| 112 | Nociti (2011) | | No | No | |
| 113 | Nociti (2011) | | No | No | |
| 114 | O'Rourke (2006) | .5 | No | No | |
| 115 | Olgiati (2016) | .4 | No | No | No |
| 116 | Panda (2014) | .01 | Yes | No | |
| 117 | Pandey (2014) | | No | No | |
| 118 | Persing (1990) | | No | No | |
| 119 | Pettigrew (1985) | 26 | No | No | |
| 120 | Plant (1989) | 52 | No | No | Yes |
| 121 | Powers (1985) | | No | No | |
| 122 | Rana (2014) | | No | No | No |
| 123 | Riley (1996) | | No | No | |
| 124 | Sabrie (2015) | .1 | No | No | |
| 125 | Salerno (1993) | 13 | No | No | No |
| 126 | Schulze-Bonhage (1995) | | No | No | |
| 127 | Schwartz (1995) | | No | No | |
| 128 | Seet (2005) | | No | Yes | No |
| 129 | Shen (2016) | 2 | No | No | |
| 130 | Singer (1998) | | No | Yes | |
| 131 | Takahashi (2009) | 52 | No | No | No |
| 132 | Tamburin (2005) | | No | No | |
| 133 | Tan (2005) | .01 | No | No | |
| 134 | Tranchant (1991) | | NR | NR | |

| | | | | | |
|---|---|---|---|---|---|
| 135 | Trankle (1997) | 17 | NR | NR | |
| 136 | Trompetto (2012) | 20 | No | No | |
| 137 | Trompetto (2012) | 12 | No | No | |
| 138 | Trompetto (2012) | 24 | Yes | No | |
| 139 | Trompetto (2012) | 104 | Yes | No | |
| 140 | Usmani (2011) | 65 | No | No | |
| 141 | Verghese (1999) | | No | No | |
| 142 | Vidailhet (1999) | 9 | No | No | |
| 143 | Vidailhet (1999) | .01 | No | No | |
| 144 | Vidailhet (1999) | .01 | No | No | |
| 145 | Vidailhet (1999) | 22 | Yes | Yes | |
| 146 | Vidailhet (1999) | 7 | No | Yes | |
| 147 | Wali (2001) | 4 | No | No | |
| 148 | Walker (2015) | 17 | No | No | No |
| 149 | Wolz (2008) | .3 | No | No | No |
| 150 | Wu (1992) | 8 | Yes | No | No |
| 151 | Zadro (2008) | .2 | No | No | |
| 152 | Garbin Di Luca (2019) | .5 | No | No | No |
| 153 | Pandey (2018) | .4 | No | No | No |
| 154 | Pandey (2018) | 20 | Yes | No | No |
| 155 | Pandey (2018) | 16 | Yes | No | No |
| 156 | Shin (2019) | 260 | No | No | No |
| 157 | Szejko (2020) | .01 | No | No | No |
| 158 | Hamed (2020) | | No | No | No |
| 159 | Li (2019) | 26 | No | No | No |
| 160 | Li (2019) | 52 | No | No | No |
| 161 | Li (2019) | 78 | Yes | No | No |
| 162 | Sul (2019) | 12 | No | No | No |
| 163 | Ghosh (2020) | | No | No | NR |
| 164 | Guimarães Rocha (2021) | 26 | Yes | No | NR |
| 165 | Guimarães Rocha (2021) | | No | No | NR |
| 166 | Han (2020) | | No | No | NR |
| 167 | Meloni (2021) | 52 | Yes | No | NR |
| 168 | Owen (2022) | 104 | No | No | NR |

| | | | | | |
|---|---|---|---|---|---|
| 169 | Umeh (2012) | 13 | No | No | NR |
| 170 | Wu, Zheng (2020) | | NR | NR | NR |
| 171 | Wu, Su (2020) | | Yes | No | NR |
| 172 | Xu (2020) | | No | No | NR |
| 173 | Yu (2021) | | Yes | No | NR |
| 174 | Aldosary (2021) | | No | No | NR |
| 175 | Smith (2021) | | No | No | NR |
| 176 | Funabe (2014) | | No | No | NR |
| 177 | Kim, Lee (2007) | | Yes | No | Yes |
| 178 | Walker (2007) | 21 | Yes | Yes | NR |
| 179 | Trompetto (2012) | 28 | Yes | No | |

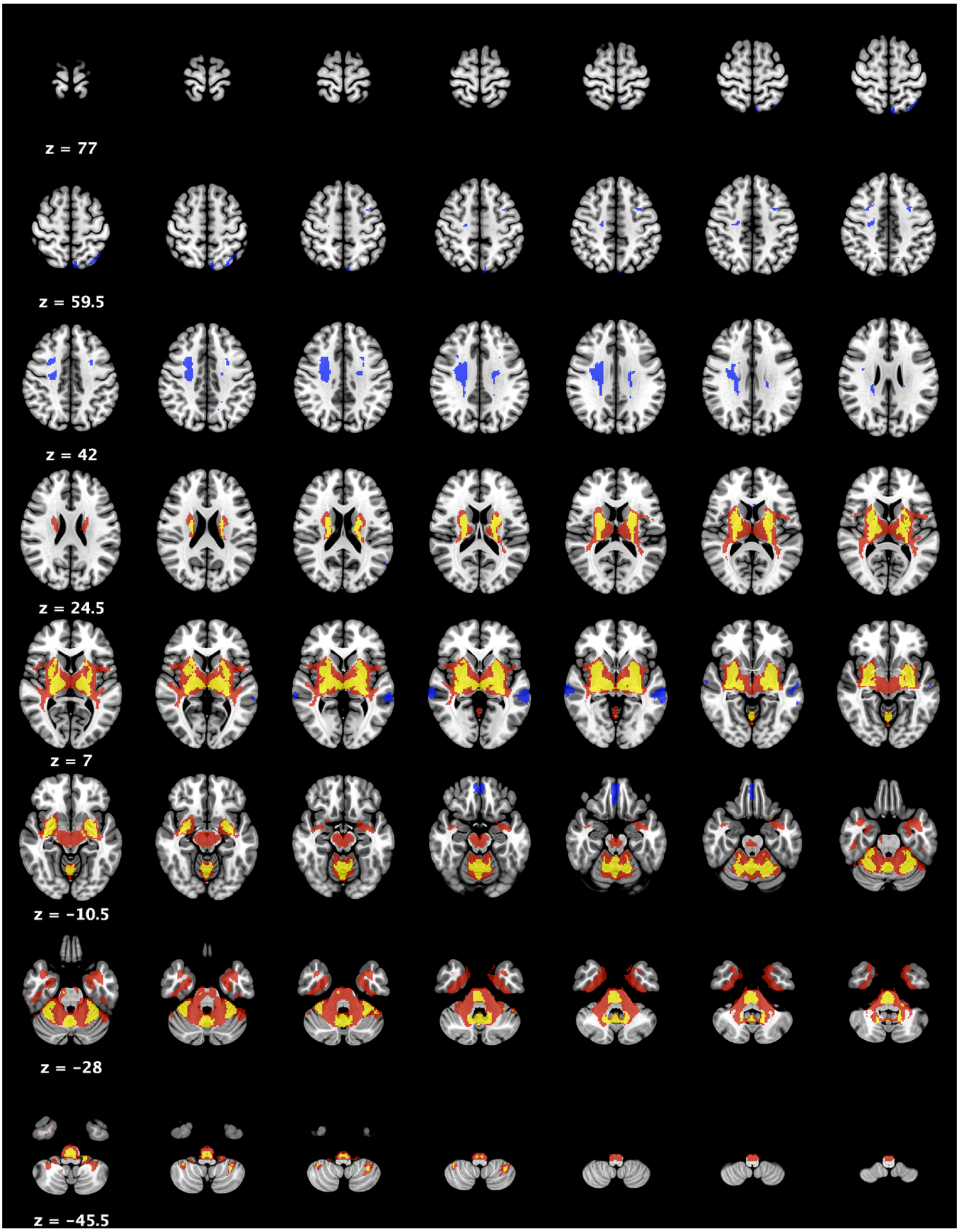

**Supplementary File 5. General dystonia network.** Axial slices showing brain regions surviving ≥3 of the four statistical tests used to define the general dystonia network. Red = 3/4 tests; yellow = 4/4 tests (positive connectivity). Blue = 3/4 tests (negative connectivity). No negatively connected voxels survived all 4 tests.

**Supplementary File 6. Clusters of the general dystonia network.** Max X, Y, Z refer to the location where the highest N of lesions were connected within the cluster, while COG X, Y, Z refers to its centre of gravity. Positively connected clusters surviving 4/4 statistical tests are shown. While no negatively connected clusters survived 4/4 tests, therefore clusters surviving 3/4 tests are shown.

**Positive connectivity**

| | N voxels | MAX X (MNI) | MAX Y | MAX Z | COG X (MNI) | COG Y | COG Z |
|---|---|---|---|---|---|---|---|
| R basal ganglia, thalamus, claustrum | 2308 | 18 | -10 | 4 | 23 | -8 | 3 |
| L basal ganglia, thalamus, claustrum | 2350 | -16 | -22 | 8 | -23 | -10 | 3 |
| Cerebellum | 2458 | -34 | -44 | -30 | -2 | -55 | -26 |
| Brainstem | 411 | 4 | -30 | -46 | 2 | -28 | -41 |

**Negative connectivity**

| | N voxels | MAX X (MNI) | MAX Y | MAX Z | COG X (MNI) | COG Y | COG Z |
|---|---|---|---|---|---|---|---|
| R subcortical WM | 792 | 22 | 0 | 32 | 25 | -10 | 36 |
| L mid/superior termporal gyrus | 286 | -66 | -30 | -2 | -61 | -31 | -2 |
| Medial frontal gyrus | 146 | -2 | 50 | -20 | 1 | 48 | -20 |
| R mid/superior termporal gyrus | 106 | 66 | -28 | -2 | 65 | -26 | -1 |
| L subcortical WM | 98 | -24 | -8 | 34 | -22 | -14 | 34 |
| L subcortical WM/middle frontal gyrus | 72 | -38 | 6 | 54 | -30 | 6 | 44 |
| L medial parietal lobe | 71 | -10 | -76 | 60 | -7 | -74 | 58 |
| L superior parietal lobe | 52 | -34 | -64 | 60 | -33 | -65 | 60 |

***Network connectivity and extent of dystonia in the body – number of overlapping voxels***

We investigated whether lesion connectivity to this dystonia brain network could predict a greater extent dystonia in the body. Results for connectivity strength and its association to expression of dystonia in the body are shown in the main manuscript. We also investigated whether the number of overlapping voxels predicted the extent of dystonia in the body. To calculate this, individual lesion network maps were binarised at a t-value of ≥7 (i.e., the same binarisation process and t-value as in the overlap analysis, described in manuscript) and the number of binarised voxels that hit/overlapped with the general dystonia network were extracted for analysis. These values were then associated with: 1) the number of body parts affected by dystonia (Spearman rank correlation); and 2) dystonia body distribution (multiple linear regression with post-hoc pairwise tests where regression showed significance). Analyses demonstrated that the number of voxels from a case's lesion network map that hit the general dystonia network was significantly associated with the number of body parts affected by dystonia ($\rho$=0.229; p=0.002) and dystonia body distribution ($F[4,174]$=3.13; p=0.016) (Figure).

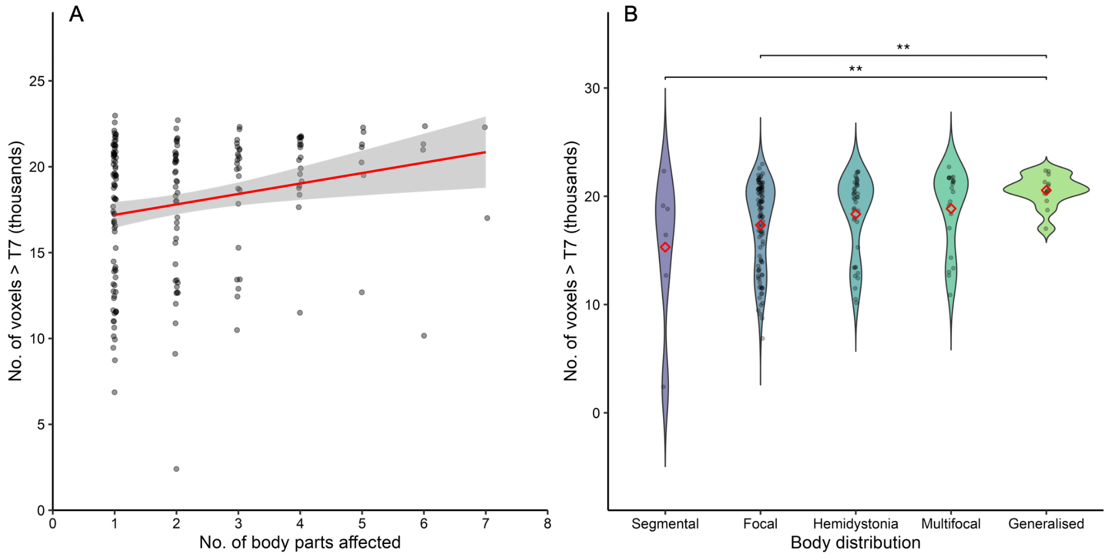

**Supplementary File 7. Network connectivity and extent of dystonia in the body.** There was a significant positive association between the number of voxels hitting the general dystonia network and the number of body parts affected by dystonia (Figure A, p=0.002). Similarly, patients with generalised dystonia had a significantly higher number of voxels hitting the general dystonia network compared to patients with focal and segmental dystonia (Figure B). ** = p<0.01. T7 = t-value of 7.

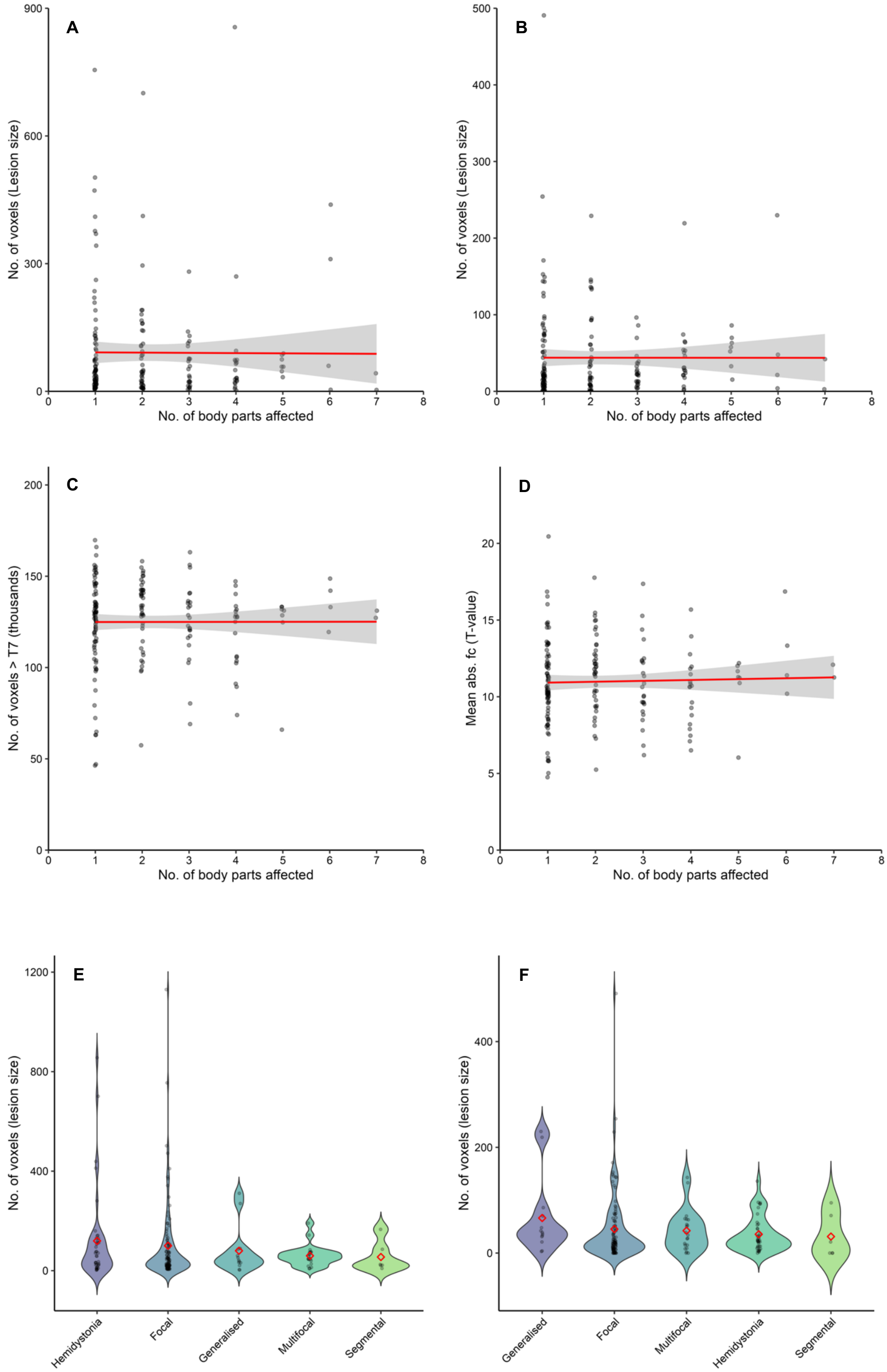
A
B
C
D
E
F
No. of voxels (Lesion size)
No. of body parts affected
No. of voxels > T7 (thousands)
Mean abs. fc (T-value)
No. of voxels (lesion size)
Hemidystonia
Focal
Generalised
Multifocal
Segmental

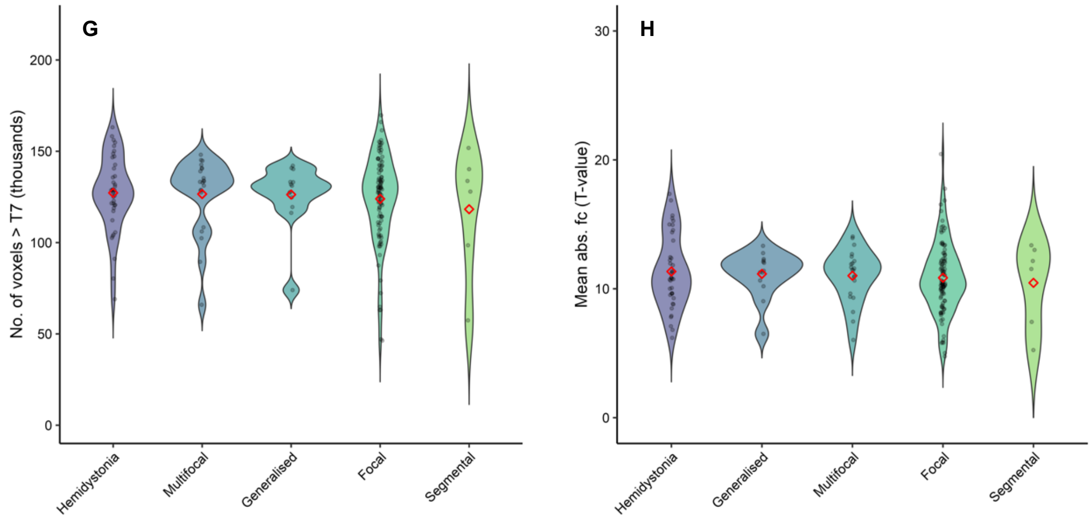

**Supplementary file 8. Network connectivity and extent of dystonia in the body.** Figure 3 in the main manuscript demonstrated that for each case's lesion network map, there were positive associations between the involvement of the general dystonia network and the extent of dystonia in the body. These analyses were not significant for lesion size or for whole brain analyses, shown here (all relationships $p>0.20$). A) (Spearman) correlation between lesion size and number of body parts affected, whole brain. B) Correlation between lesion size and number of body parts affected, general dystonia network. C) Correlation between number of voxels ≥T7 from a case's lesion network map and number of body parts affected, whole brain. D) Correlation between mean absolute functional connectivity of T-maps and number of body parts affected, whole brain. E) Regression analysis of lesion size between the dystonia body distributions, whole brain. F) Regression analysis of lesion size between the dystonia body distributions, general dystonia network. G) Regression analysis of number of voxels ≥T7 from a case's lesion network map between the dystonia body distributions, whole brain. H) Regression analysis of mean absolute functional between dystonia body distributions, whole brain.

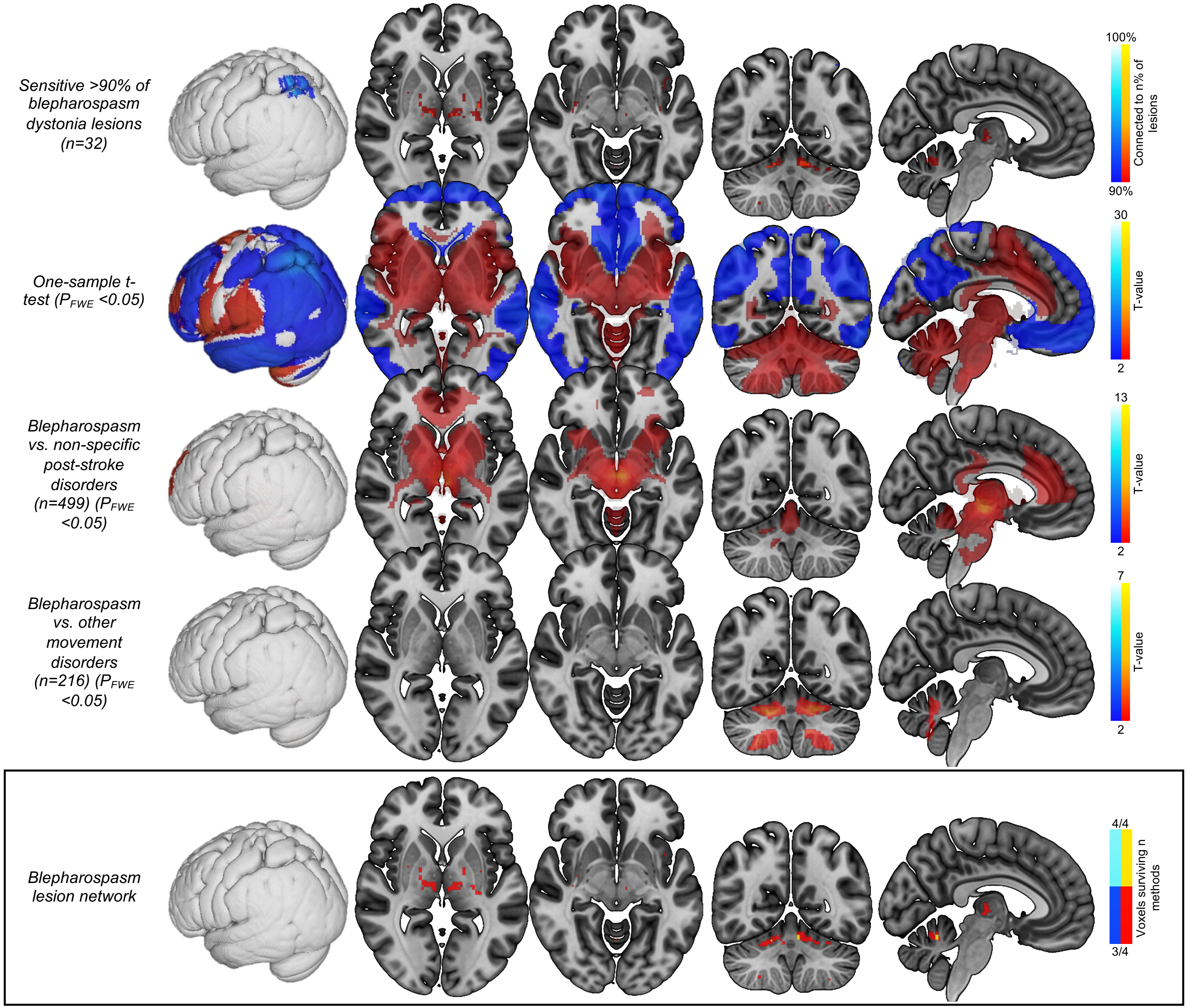

**Supplementary file 9. Body part and distribution lesion networks.** Figure shows the individual tests that were used to define each of the body part and body distribution lesion networks.

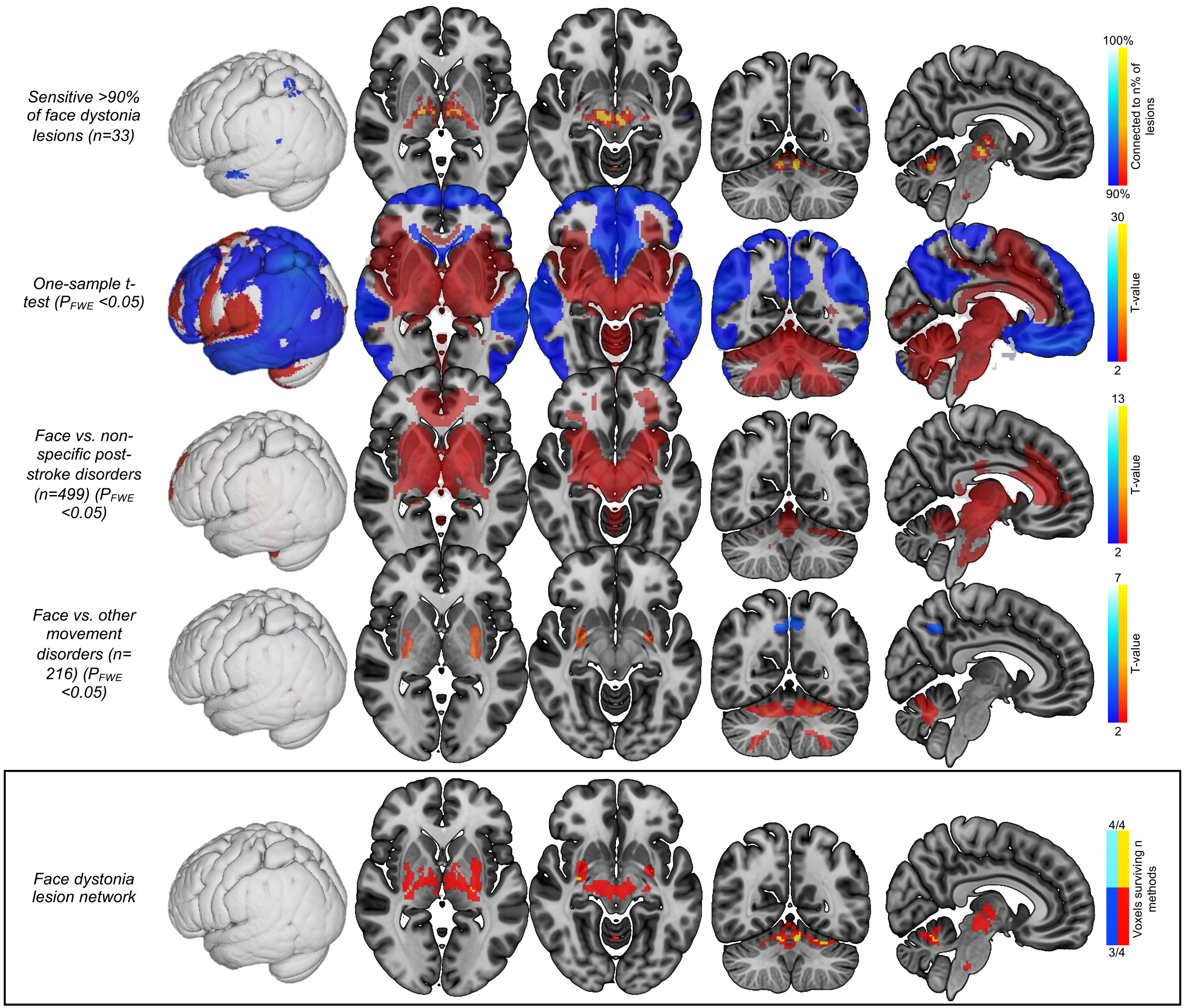

Sensitive >90% of face dystonia lesions (n=33)
One-sample t-test (PFWE <0.05)
Face vs. non-specific post-stroke disorders (n=499) (PFWE <0.05)
Face vs. other movement disorders (n= 216) (PFWE <0.05)
Face dystonia lesion network
100%
Connected to n% of lesions
90%
30
T-value
2
13
T-value
2
7
T-value
2
4/4
Voxels surviving n methods
3/4

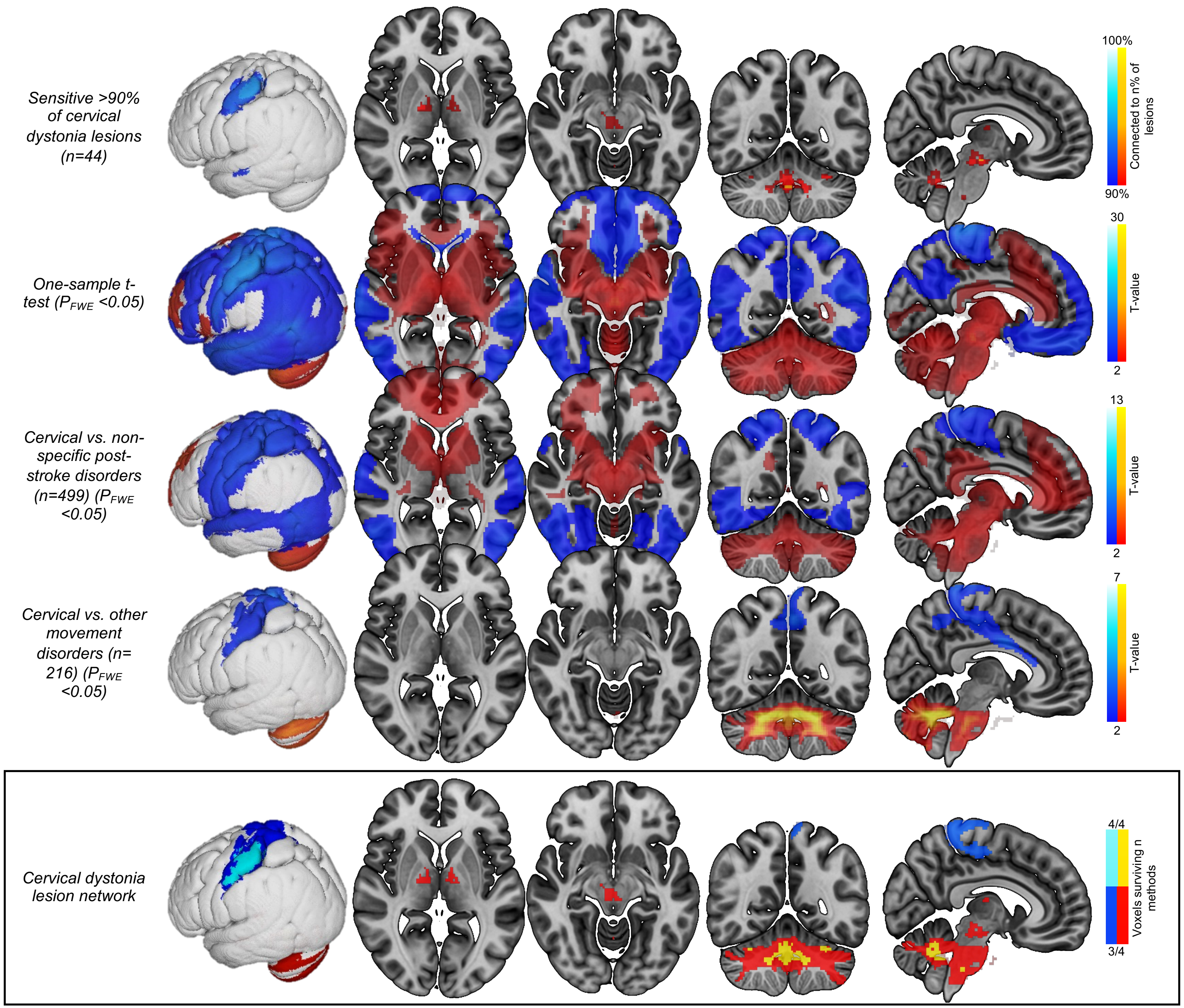

Sensitive >90% of cervical dystonia lesions (n=44)
One-sample t-test ($P_{FWE}$ <0.05)
Cervical vs. non-specific post-stroke disorders (n=499) ($P_{FWE}$ <0.05)
Cervical vs. other movement disorders (n= 216) ($P_{FWE}$ <0.05)
Cervical dystonia lesion network
100%
Connected to n% of lesions
90%
30
T-value
2
13
T-value
2
7
T-value
2
4/4
Voxels surviving n methods
3/4

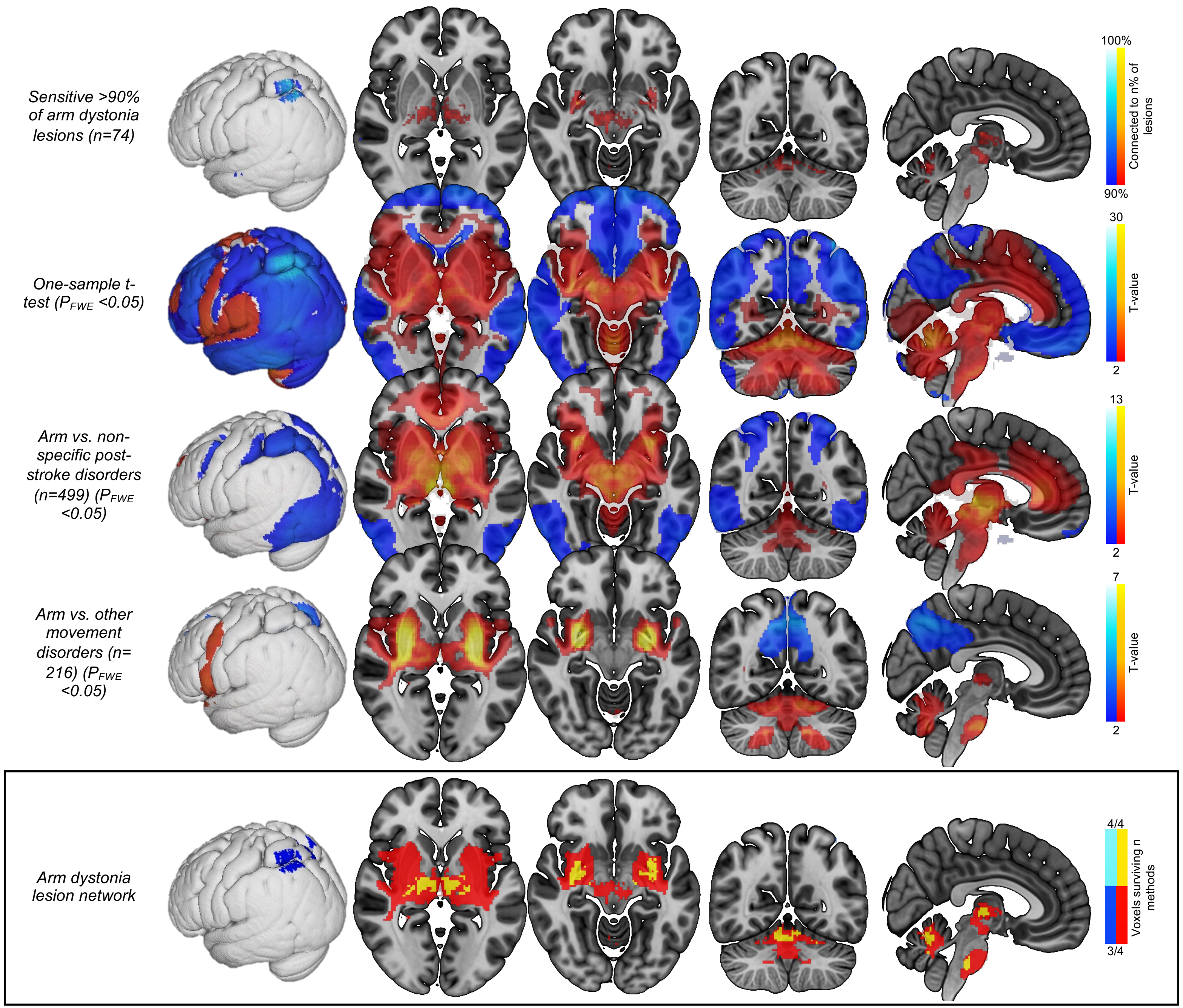

Sensitive >90% of arm dystonia lesions (n=74)
One-sample t-test (PFWE <0.05)
Arm vs. non-specific post-stroke disorders (n=499) (PFWE <0.05)
Arm vs. other movement disorders (n=216) (PFWE <0.05)
Arm dystonia lesion network
100%
Connected to n% of lesions
90%
30
T-value
2
13
T-value
2
7
T-value
2
4/4
Voxels surviving n methods
3/4

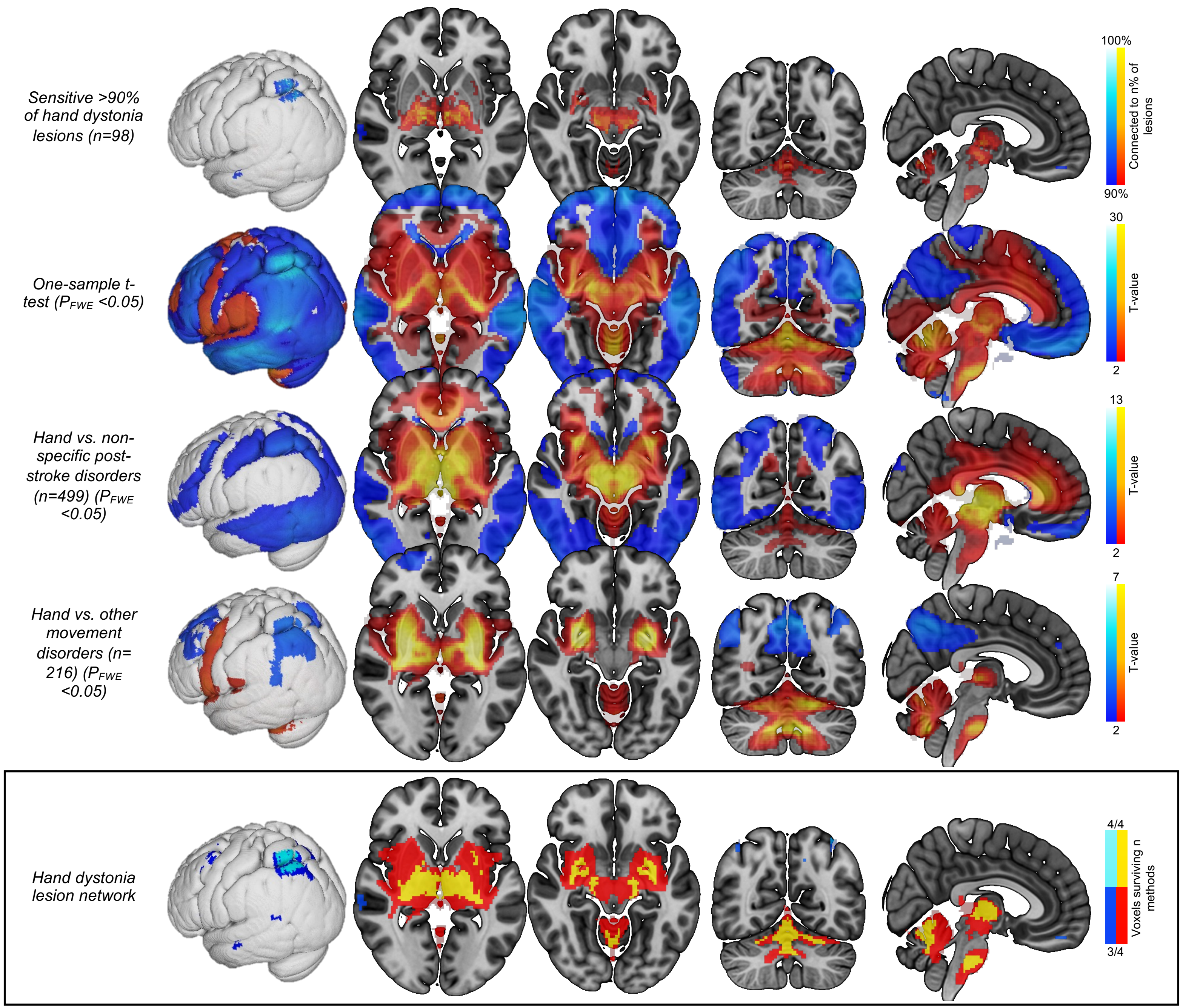

Sensitive >90% of hand dystonia lesions (n=98)
One-sample t-test ($P_{FWE}$ <0.05)
Hand vs. non-specific post-stroke disorders (n=499) ($P_{FWE}$ <0.05)
Hand vs. other movement disorders (n=216) ($P_{FWE}$ <0.05)
Hand dystonia lesion network
100%
Connected to n% of lesions
90%
30
T-value
2
13
T-value
2
7
T-value
2
4/4
Voxels surviving n methods
3/4

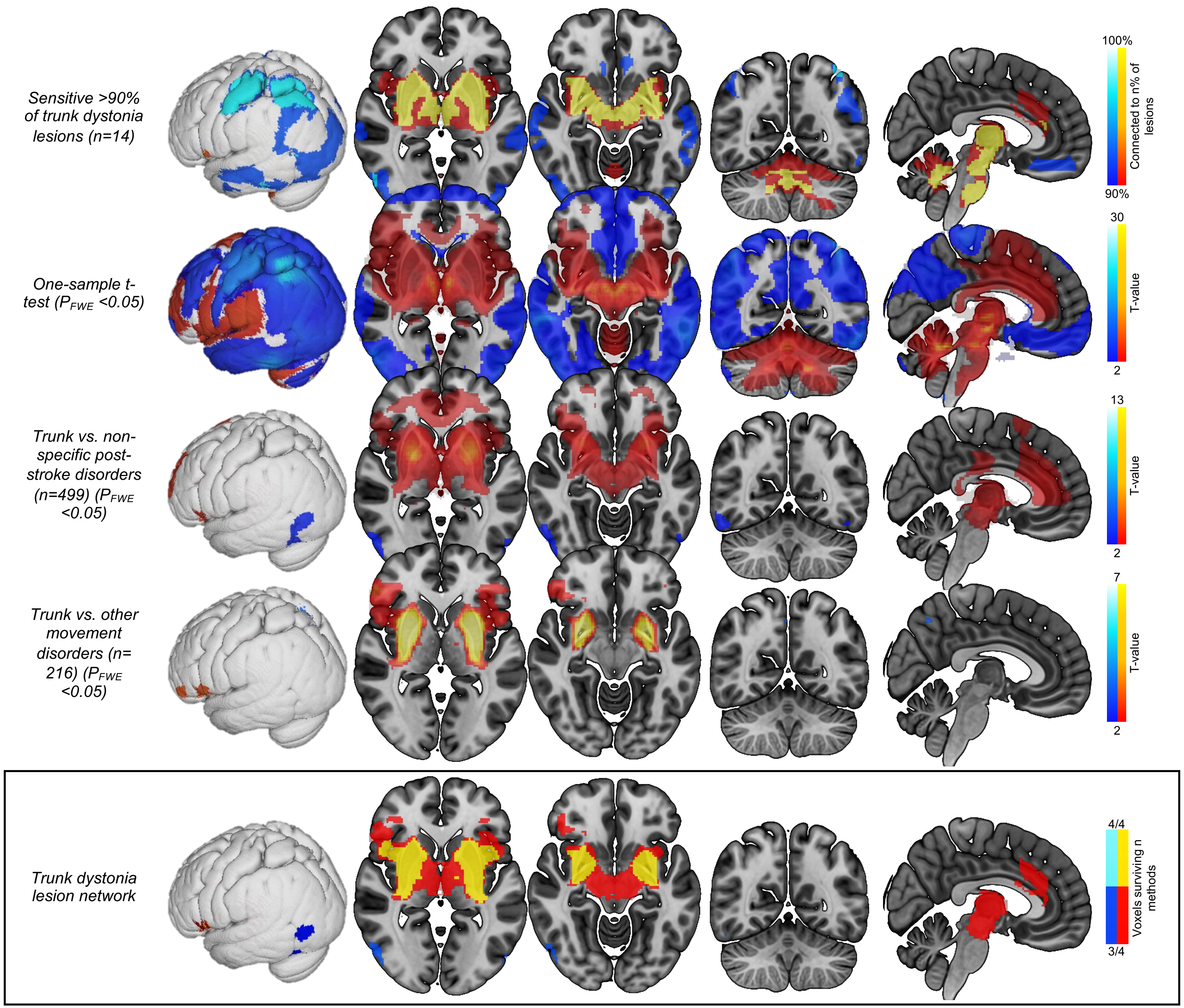

Sensitive >90% of trunk dystonia lesions (n=14)
One-sample t-test ($P_{FWE}$ <0.05)
Trunk vs. non-specific post-stroke disorders (n=499) ($P_{FWE}$ <0.05)
Trunk vs. other movement disorders (n=216) ($P_{FWE}$ <0.05)
Trunk dystonia lesion network
100%
90%
Connected to n% of lesions
30
2
T-value
13
2
T-value
7
2
T-value
4/4
3/4
Voxels surviving n methods

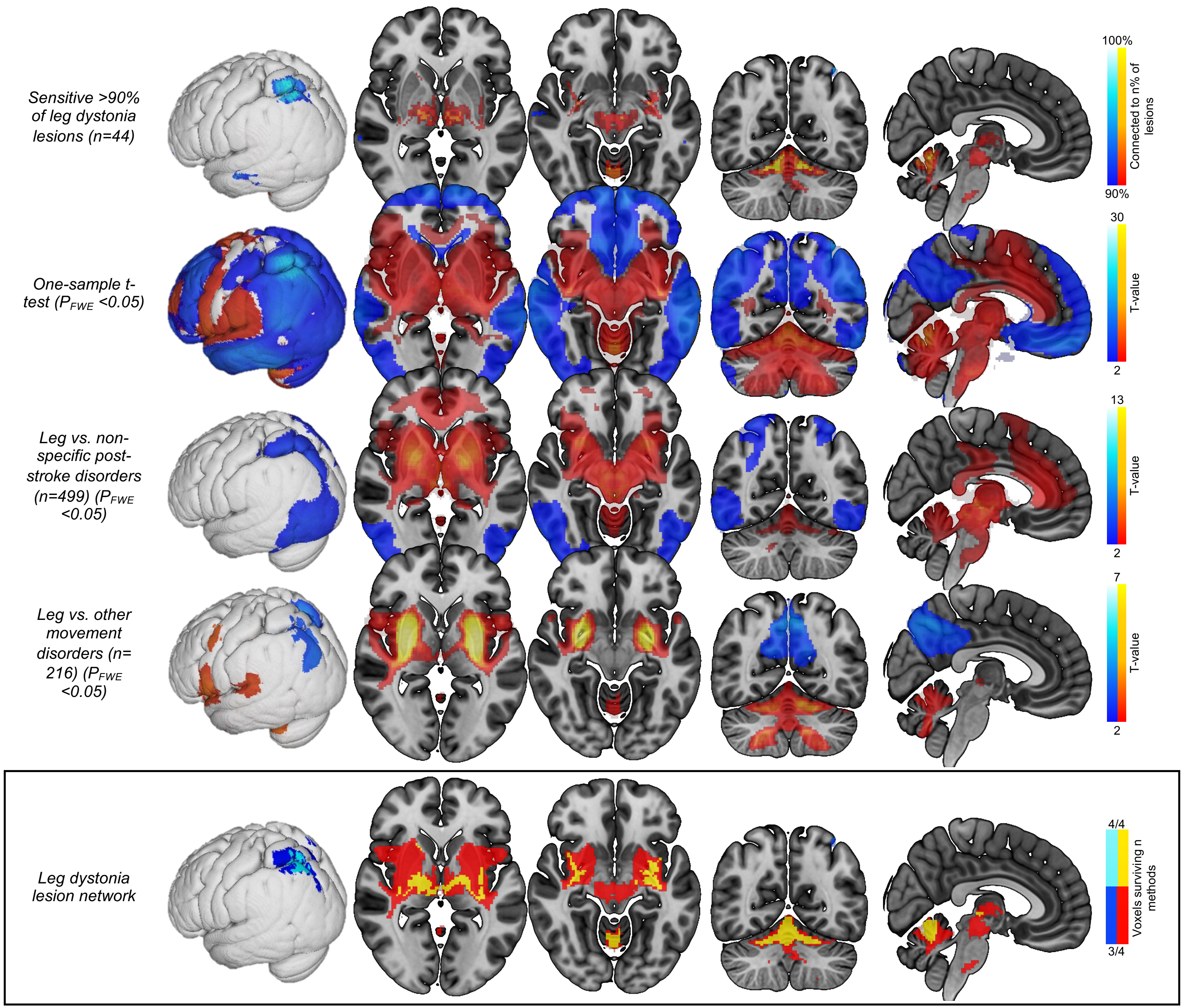
Sensitive >90% of leg dystonia lesions (n=44)
One-sample t-test ($P_{FWE}$ <0.05)
Leg vs. non-specific post-stroke disorders (n=499) ($P_{FWE}$ <0.05)
Leg vs. other movement disorders (n= 216) ($P_{FWE}$ <0.05)
Leg dystonia lesion network
100%
Connected to n% of lesions
90%
30
T-value
2
13
T-value
2
7
T-value
2
4/4
Voxels surviving n methods
3/4

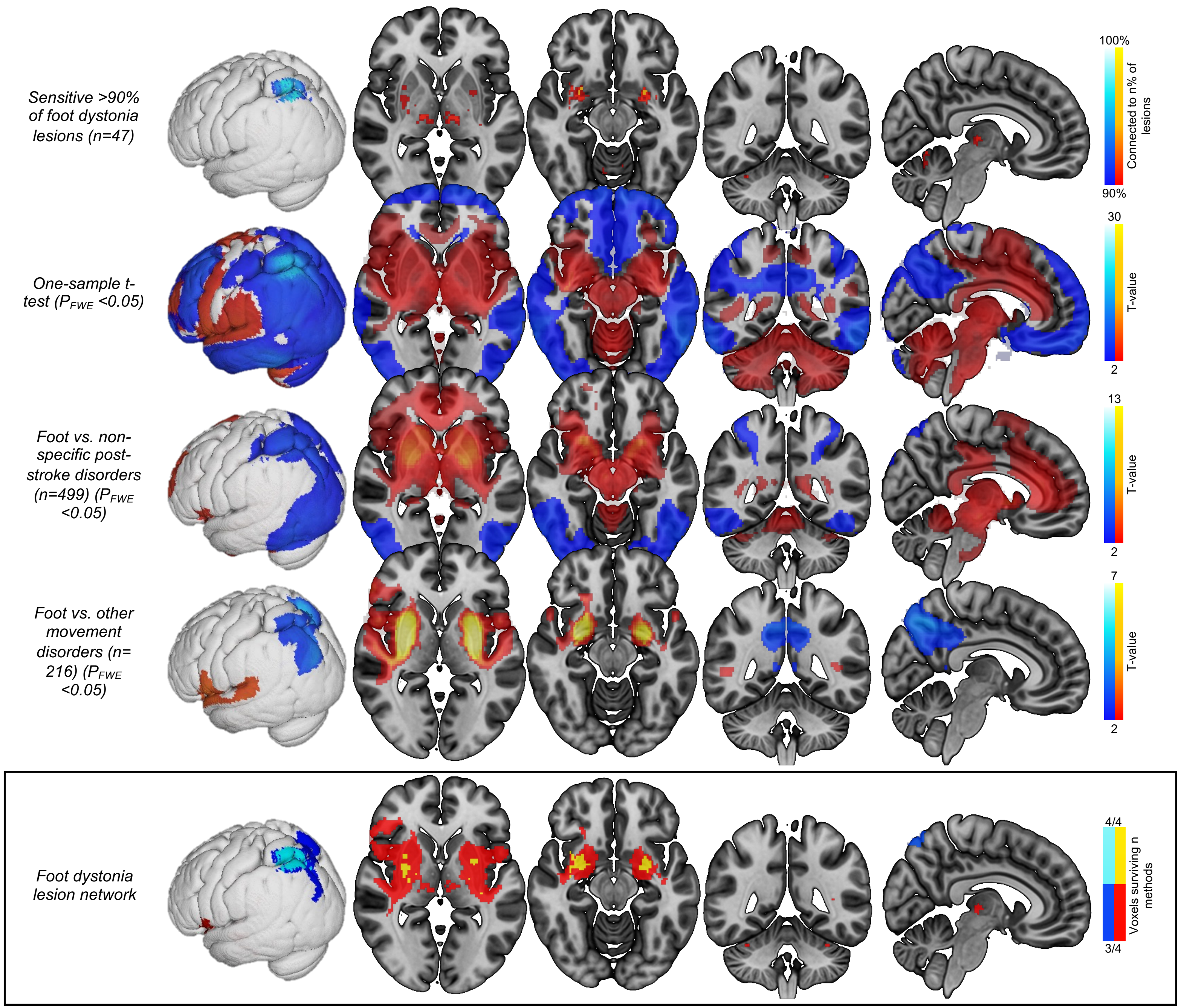

Sensitive >90% of foot dystonia lesions (n=47)
One-sample t-test ($P_{FWE}$ <0.05)
Foot vs. non-specific post-stroke disorders (n=499) ($P_{FWE}$ <0.05)
Foot vs. other movement disorders (n=216) ($P_{FWE}$ <0.05)
Foot dystonia lesion network
100%
Connected to n% of lesions
90%
30
T-value
2
13
T-value
2
7
T-value
2
4/4
Voxels surviving n methods
3/4

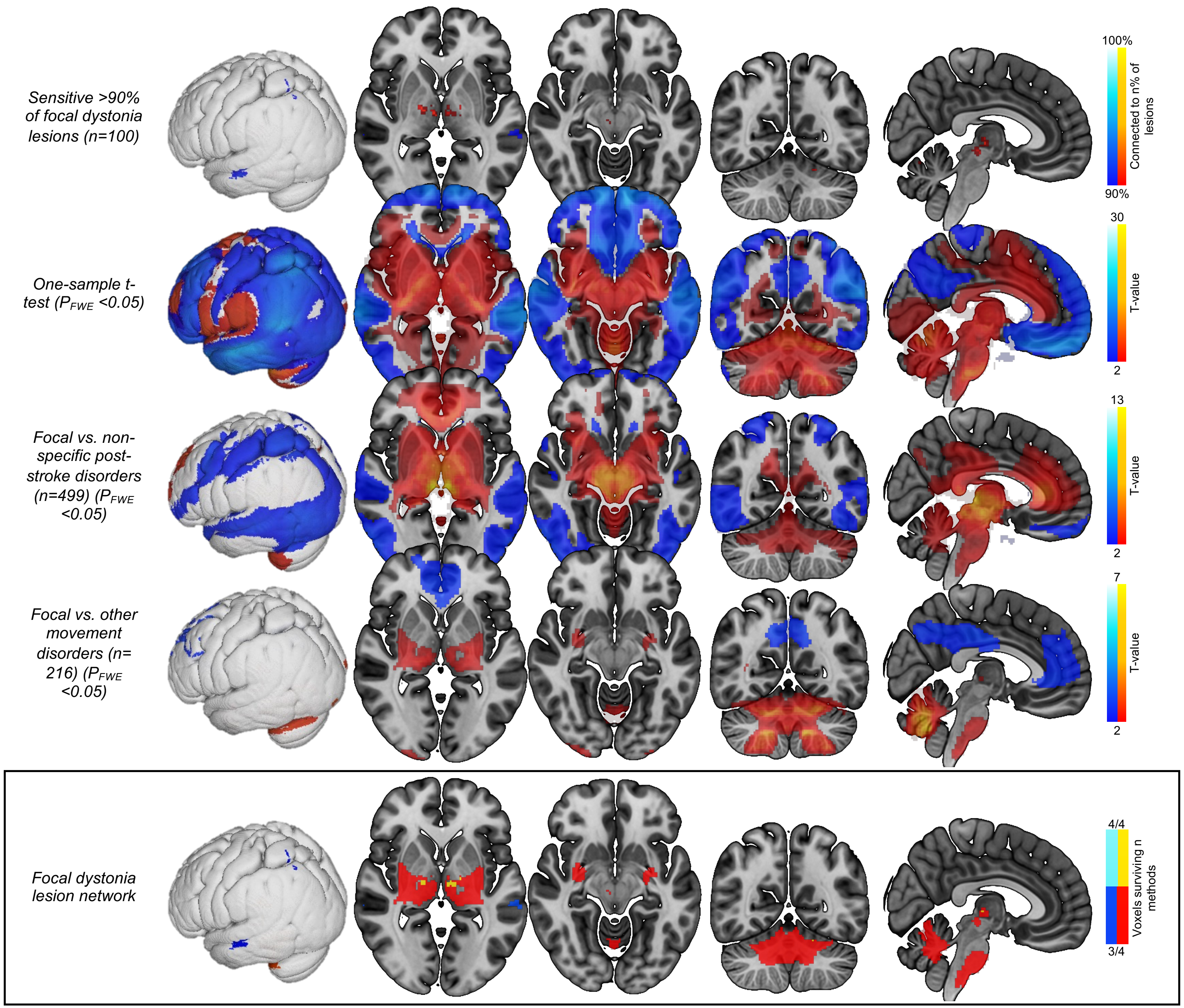

Sensitive >90% of focal dystonia lesions (n=100)
One-sample t-test ($P_{FWE}$ <0.05)
Focal vs. non-specific post-stroke disorders (n=499) ($P_{FWE}$ <0.05)
Focal vs. other movement disorders (n=216) ($P_{FWE}$ <0.05)
Focal dystonia lesion network
100%
Connected to n% of lesions
90%
30
T-value
2
13
T-value
2
7
T-value
2
4/4
Voxels surviving n methods
3/4

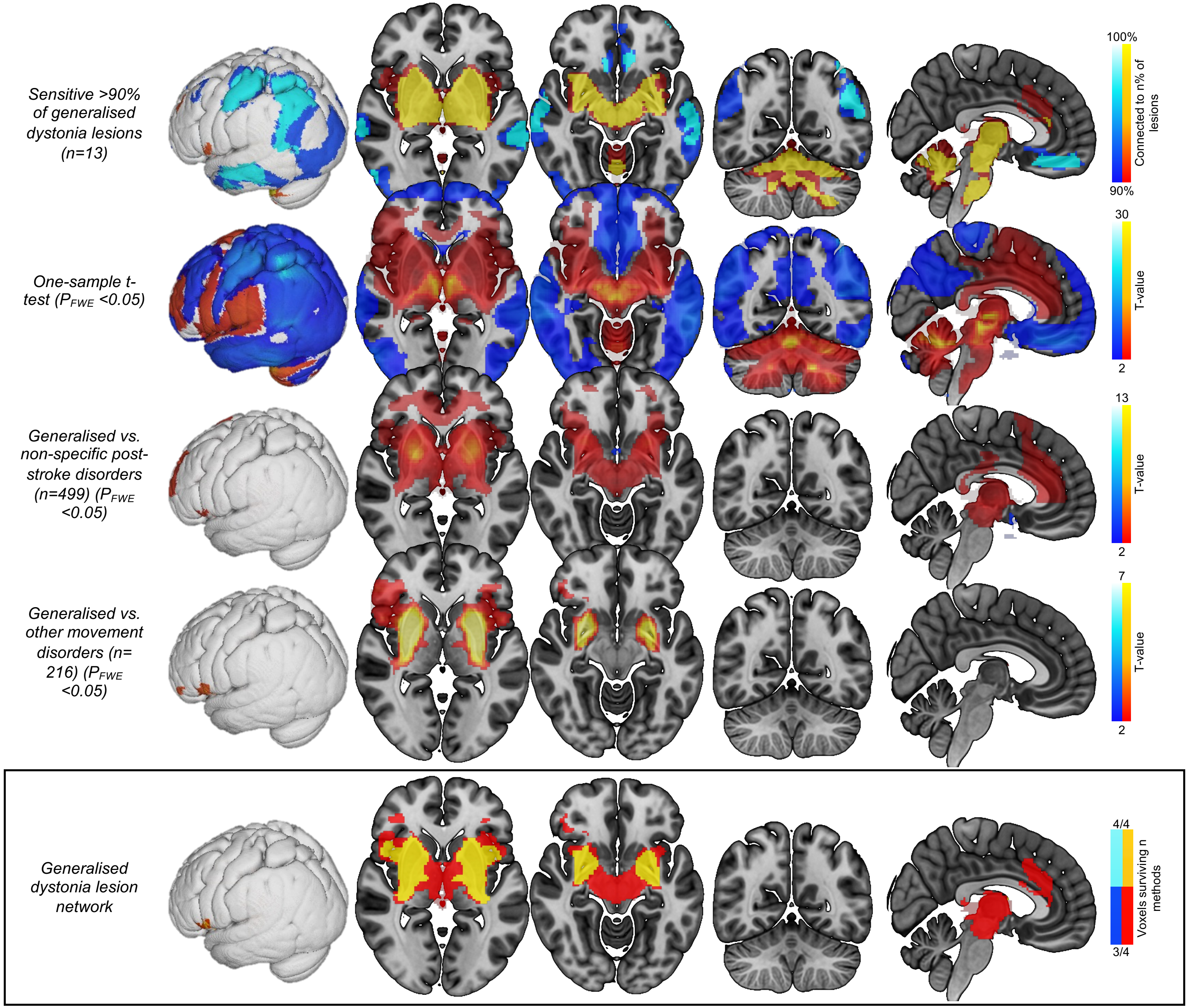

Sensitive >90% of generalised dystonia lesions (n=13)
One-sample t-test ($P_{FWE}$ <0.05)
Generalised vs. non-specific post-stroke disorders (n=499) ($P_{FWE}$ <0.05)
Generalised vs. other movement disorders (n=216) ($P_{FWE}$ <0.05)
Generalised dystonia lesion network
100%
90%
Connected to n% of lesions
30
2
T-value
13
2
T-value
7
2
T-value
4/4
3/4
Voxels surviving n methods

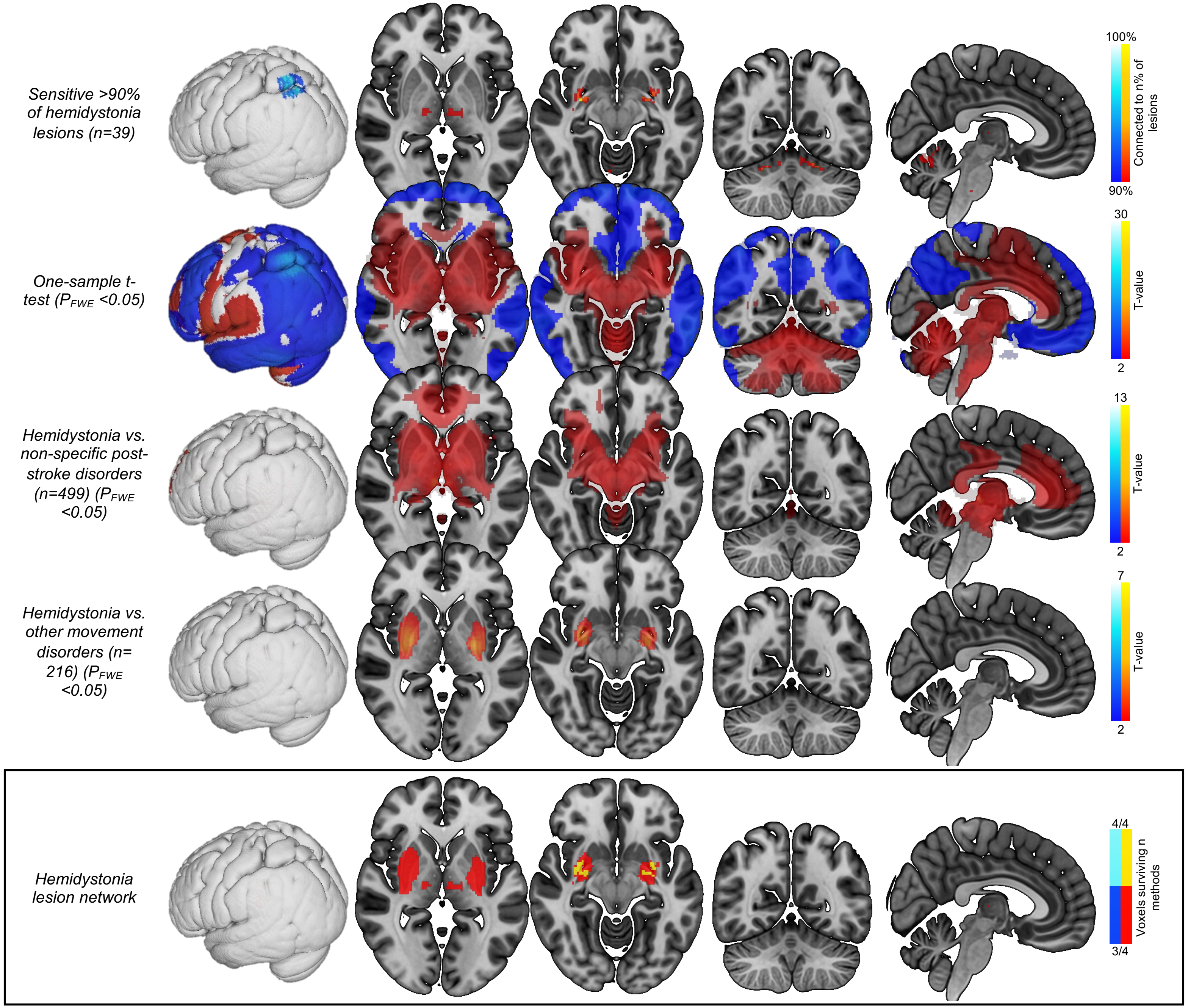

Sensitive >90% of hemidystonia lesions (n=39)
One-sample t-test ($P_{FWE}$ <0.05)
Hemidystonia vs. non-specific post-stroke disorders (n=499) ($P_{FWE}$ <0.05)
Hemidystonia vs. other movement disorders (n=216) ($P_{FWE}$ <0.05)
Hemidystonia lesion network
100%
Connected to n% of lesions
90%
30
T-value
2
13
T-value
2
7
T-value
2
4/4
Voxels surviving n methods
3/4

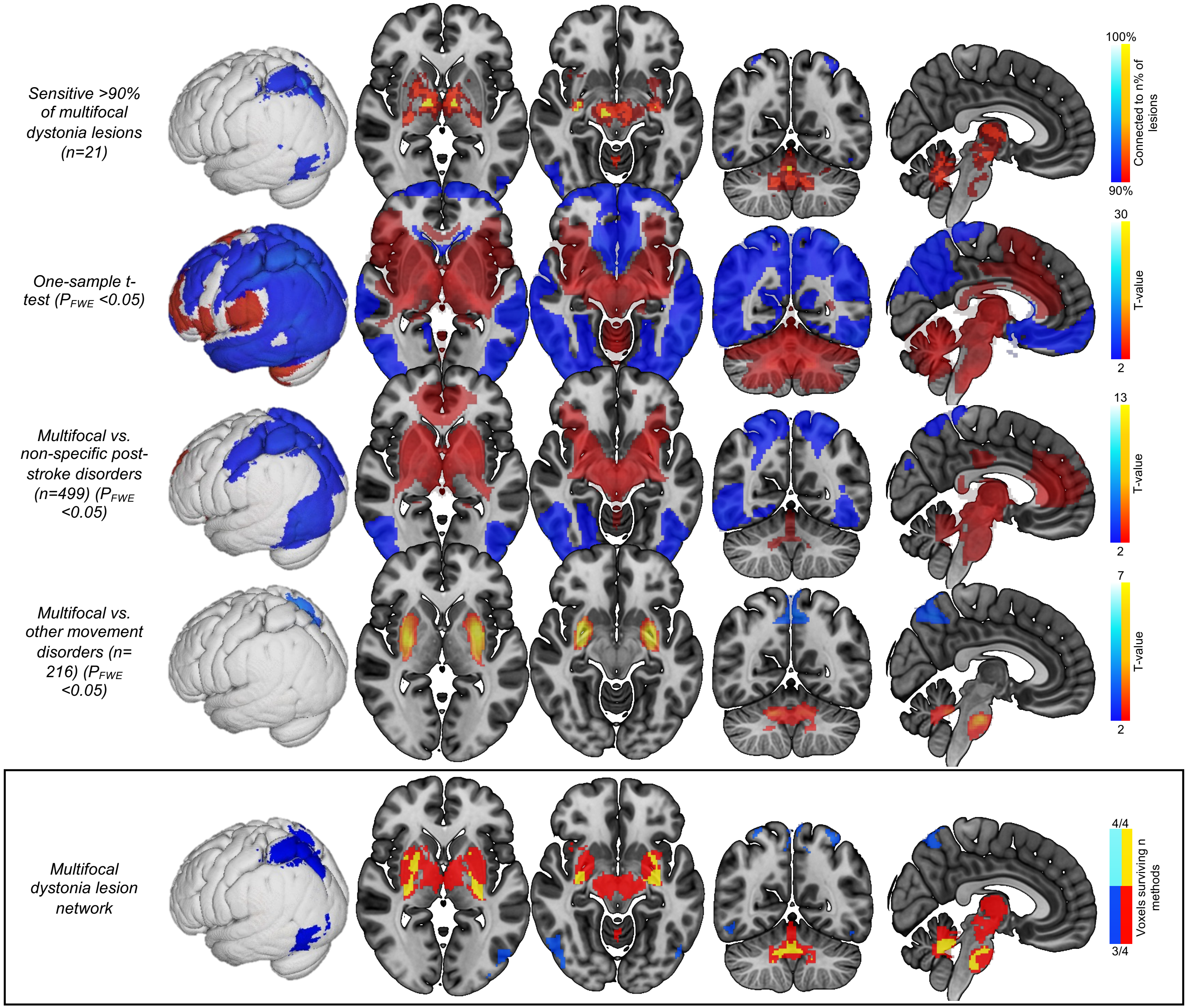

Sensitive >90% of multifocal dystonia lesions (n=21)
One-sample t-test ($P_{FWE}$ <0.05)
Multifocal vs. non-specific post-stroke disorders (n=499) ($P_{FWE}$ <0.05)
Multifocal vs. other movement disorders (n=216) ($P_{FWE}$ <0.05)
Multifocal dystonia lesion network
100%
Connected to n% of lesions
90%
30
T-value
2
13
T-value
2
7
T-value
2
4/4
Voxels surviving n methods
3/4

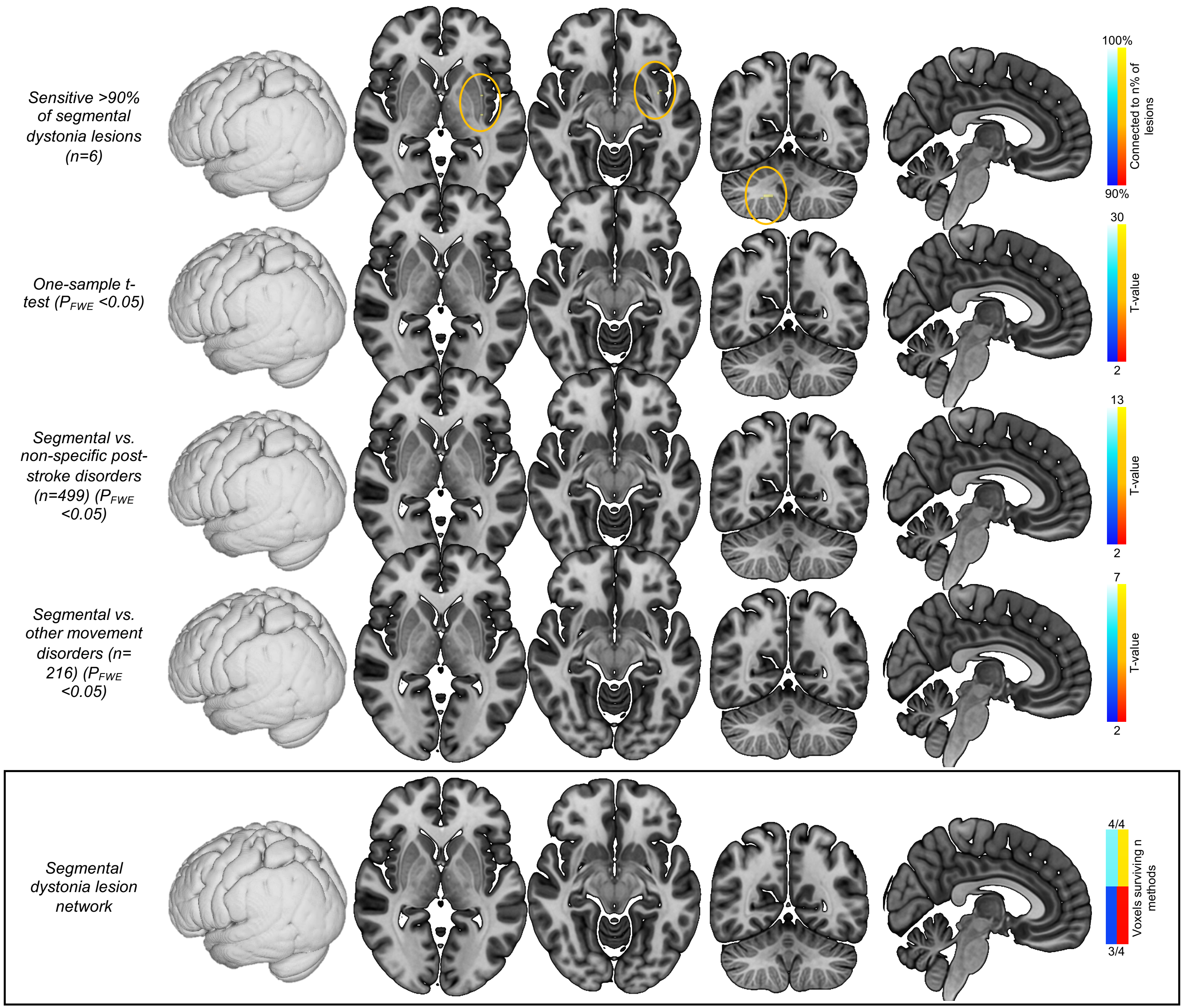

Sensitive >90% of segmental dystonia lesions (n=6)
One-sample t-test ($P_{FWE}$ <0.05)
Segmental vs. non-specific post-stroke disorders (n=499) ($P_{FWE}$ <0.05)
Segmental vs. other movement disorders (n= 216) ($P_{FWE}$ <0.05)
Segmental dystonia lesion network
100%
Connected to n% of lesions
90%
30
T-value
2
13
T-value
2
7
T-value
2
4/4
Voxels surviving n methods
3/4

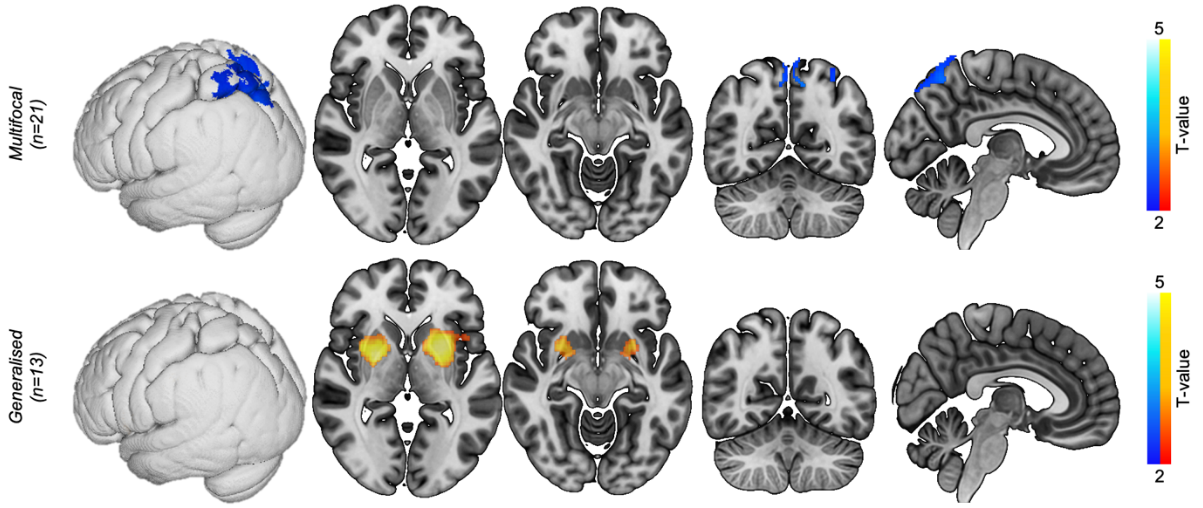

**Supplementary File 10. Significant differences between dystonias - body distribution.** We investigated whether there were significant differences in lesion connectivity between body distributions. Here, each dystonia body distribution was compared to other dystonia cases without that that body distribution. The generalised and trunk dystonia findings were similar given that 13/14 cases with trunk dystonia were categorised as having generalised dystonia. T-values are shown for visual purposes, masked to regions p<0.05 FWE corrected. Body distributions not shown did not demonstrate significant differences.

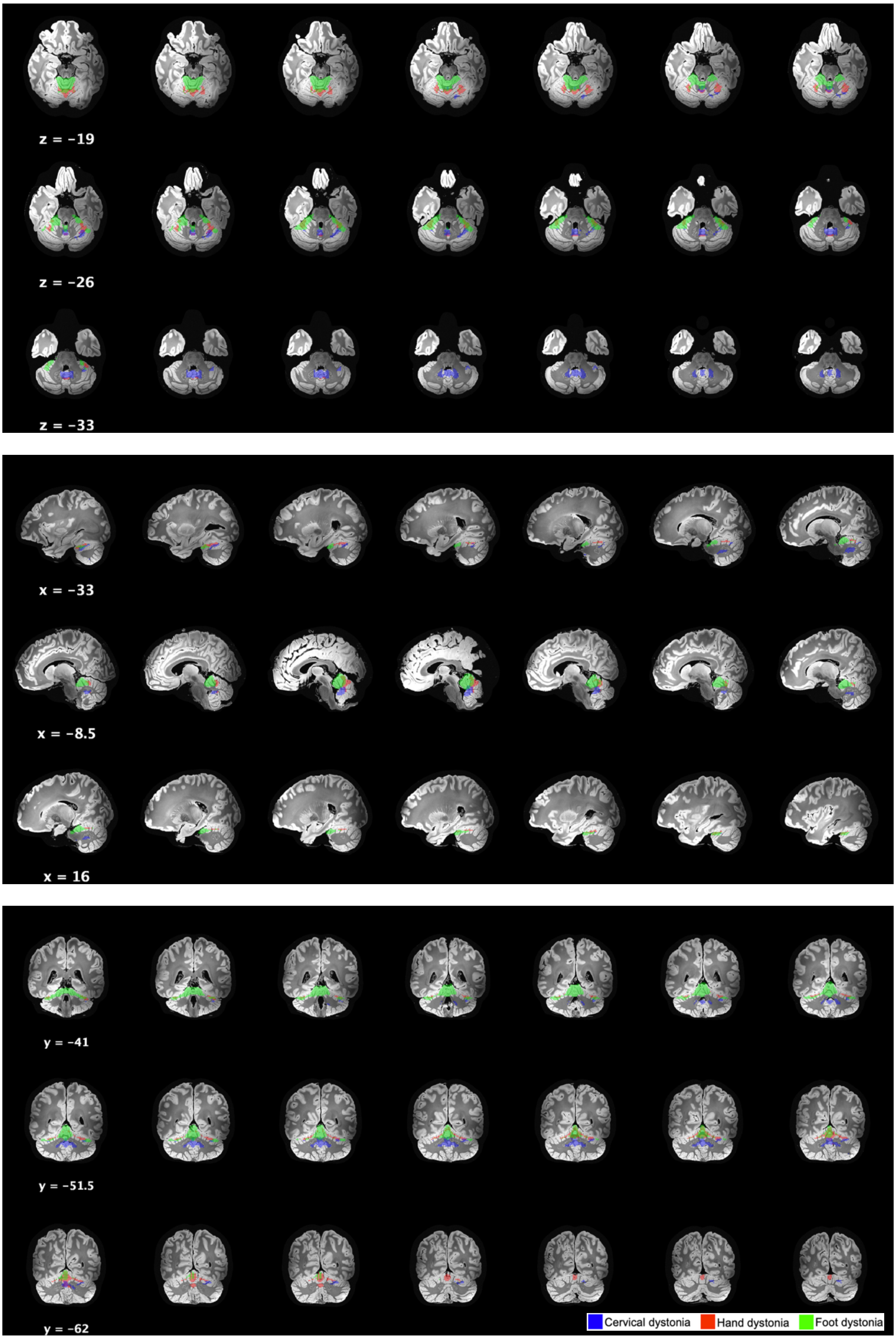

**Supplementary File 11.** Additional slices of cervical, hand, and foot dystonia lesion networks in the cerebellum.

**Supplementary File 12.** Number of voxels of the cervical, hand, and foot dystonia network falling within the sub-structures of the SUIT atlas of the cerebellum. Sub-structures not shown did not contain at least 10 network voxels or have voxels cover at least 10% of the atlas region.

| <u>**Cervical dystonia**</u> | ***Network voxels*** | ***Total voxels of structure*** | ***% of atlas region implicated*** |
|---|---|---|---|
| Left VI | 147 | 1561 | 9.4 |
| Vermis IX | 81 | 128 | 63.3 |
| Left Dentate | 76 | 250 | 30.4 |
| Right White | 38 | 1018 | 3.7 |
| Left IX | 34 | 604 | 5.6 |
| Vermis X | 33 | 59 | 55.9 |
| Right Dentate | 29 | 255 | 11.4 |
| Left White | 28 | 1069 | 2.6 |
| Right IX | 27 | 593 | 4.6 |
| Vermis VIIIa | 18 | 194 | 9.3 |
| Vermis VI | 17 | 358 | 4.7 |
| Left Interposed | 11 | 38 | 28.9 |
| Right Interposed | 10 | 38 | 26.3 |
| Right Fastigial | 6 | 7 | 85.7 |
| Left Fastigial | 5 | 6 | 83.3 |
| <u>**Hand dystonia**</u> | | | |
| Left VI | 219 | 1561 | 14.0 |
| Right V | 167 | 738 | 22.6 |
| Left V | 164 | 779 | 21.1 |
| Right VI | 110 | 1417 | 7.8 |
| Vermis VI | 83 | 358 | 23.2 |
| Vermis VIIIa | 22 | 194 | 11.3 |
| Left I IV | 17 | 632 | 2.7 |
| Right I IV | 13 | 663 | 2.0 |
| <u>**Foot dystonia**</u> | | | |
| Right I IV | 582 | 663 | 87.8 |
| Left I IV | 515 | 632 | 81.5 |
| Right V | 277 | 738 | 37.5 |
| Left V | 230 | 779 | 29.5 |
| Right VI | 151 | 1417 | 10.7 |
| Left VI | 106 | 1561 | 6.8 |
| Left White | 37 | 1069 | 3.5 |
| Left CrusI | 35 | 2210 | 1.6 |
| Right CrusI | 25 | 2230 | 1.1 |
| Right White | 24 | 1018 | 2.4 |
| Right Fastigial | 3 | 7 | 42.9 |
| Left Fastigial | 2 | 6 | 33.3 |

**Supplementary File 13. Dystonia lesion network maps in the thalamus and putamen.** Our primary analysis demonstrated that cervical, hand, and foot dystonia localised different sub-regions of the cerebellum (Figure 6, main manuscript). To test whether these findings were also present within other brain regions, we visualised these cervical, hand, and foot dystonia lesion network maps within the thalamus and the putamen, two other strongly localised structures in our analyses. Similar methods were used as in the cerebellar analysis. Specifically, cervical, hand, and foot lesion networks (Figure 4, main manuscript) were overlaid onto a standard adult brain, masked to the thalamus MNI structural probabilistic atlas, 25% probability) and putamen (Harvard-Oxford probabilistic subcortical atlas, 25% probability, and compared visually. As with the cerebellum analysis, the thresholds of these maps were adjusted to attempt to localise the major sub-region implicated, and to show a similar number of voxels between the groups, where possible. The following thresholds were used – **Thalamus** = Foot dystonia network map: voxels surviving ≥3 of 4 statistical tests, as shown in Figure 4; hand: 4/4; cervical: ≥3/4. **Putamen** = Foot: 4/4; hand: 4/4; cervical: ≥2/4.

Similar to the analysis within the cerebellum, there were differences in the sub-regions of the thalamus and putamen localised between cervical, hand, and foot dystonia (Figure). Specifically, compared to hand and foot dystonia, cervical dystonia preferentially mapped to superior and anterior portions of the thalamus, and superior and anterior portions of the putamen. Hand dystonia preferentially mapped to posterior regions of the putamen.

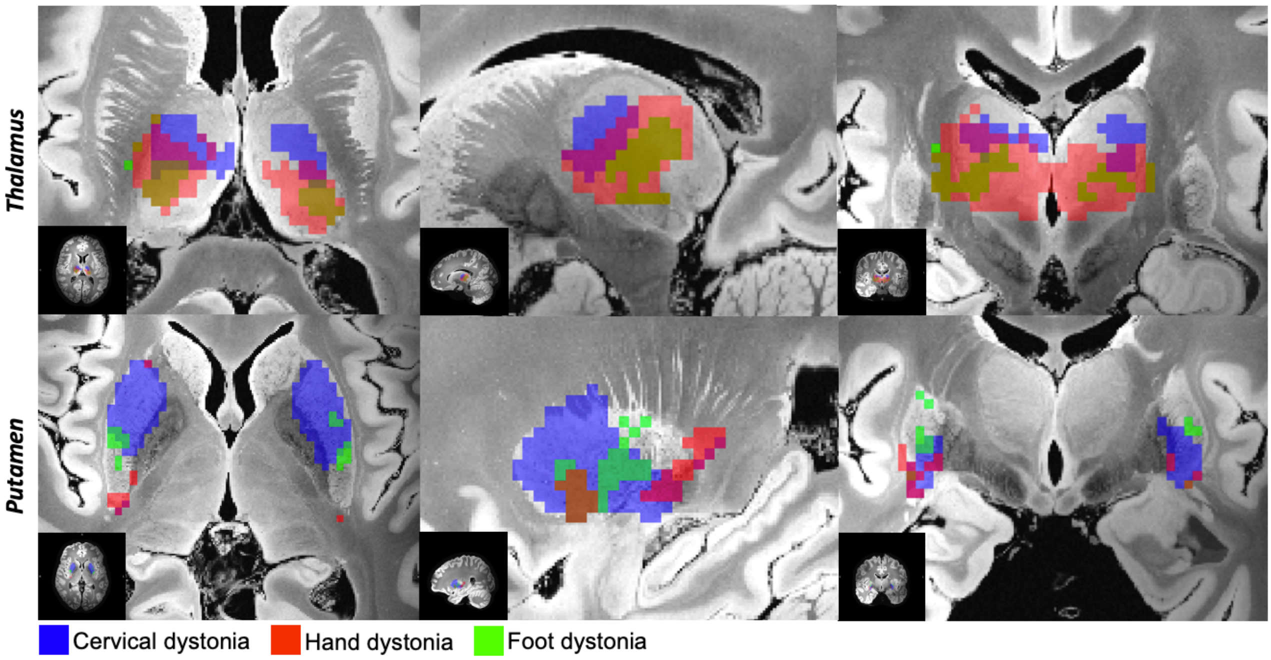

**Supplementary File 13A. Dystonia lesion network maps in the thalamus and putamen.** As with the cerebellum (Figure 7, main manuscript), cervical, hand, and foot dystonia lesion networks mapped to different sub-regions within the thalamus and putamen. Slices shown from L to R: Thalamus = Z: 11, X: -15, Y: -14.5. Putamen = Z: 1, X: 27.5, Y: -8.5. Additional slices are shown below.

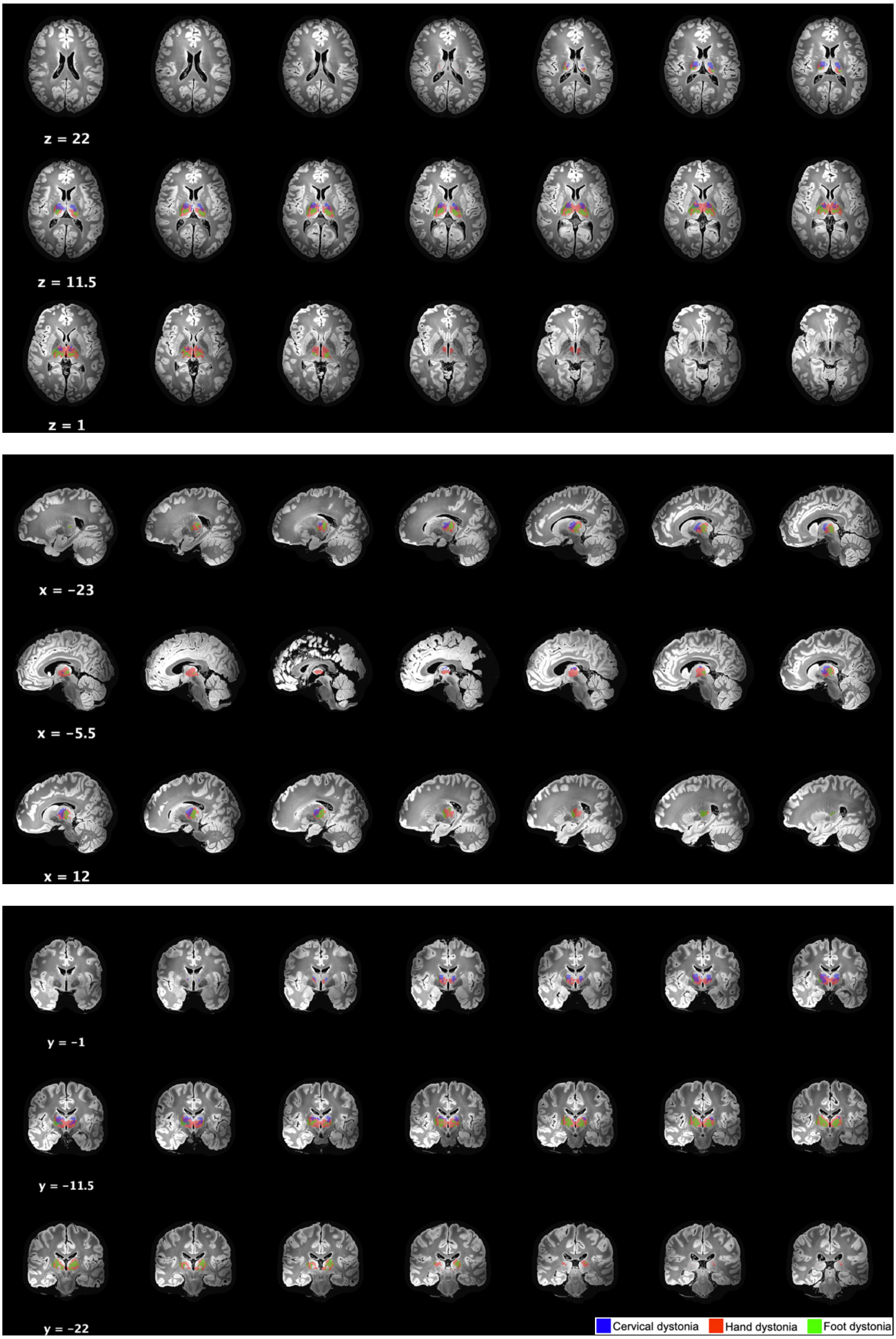

**Supplementary File 13B.** Additional slices of cervical, hand, and foot dystonia lesion networks in the thalamus.

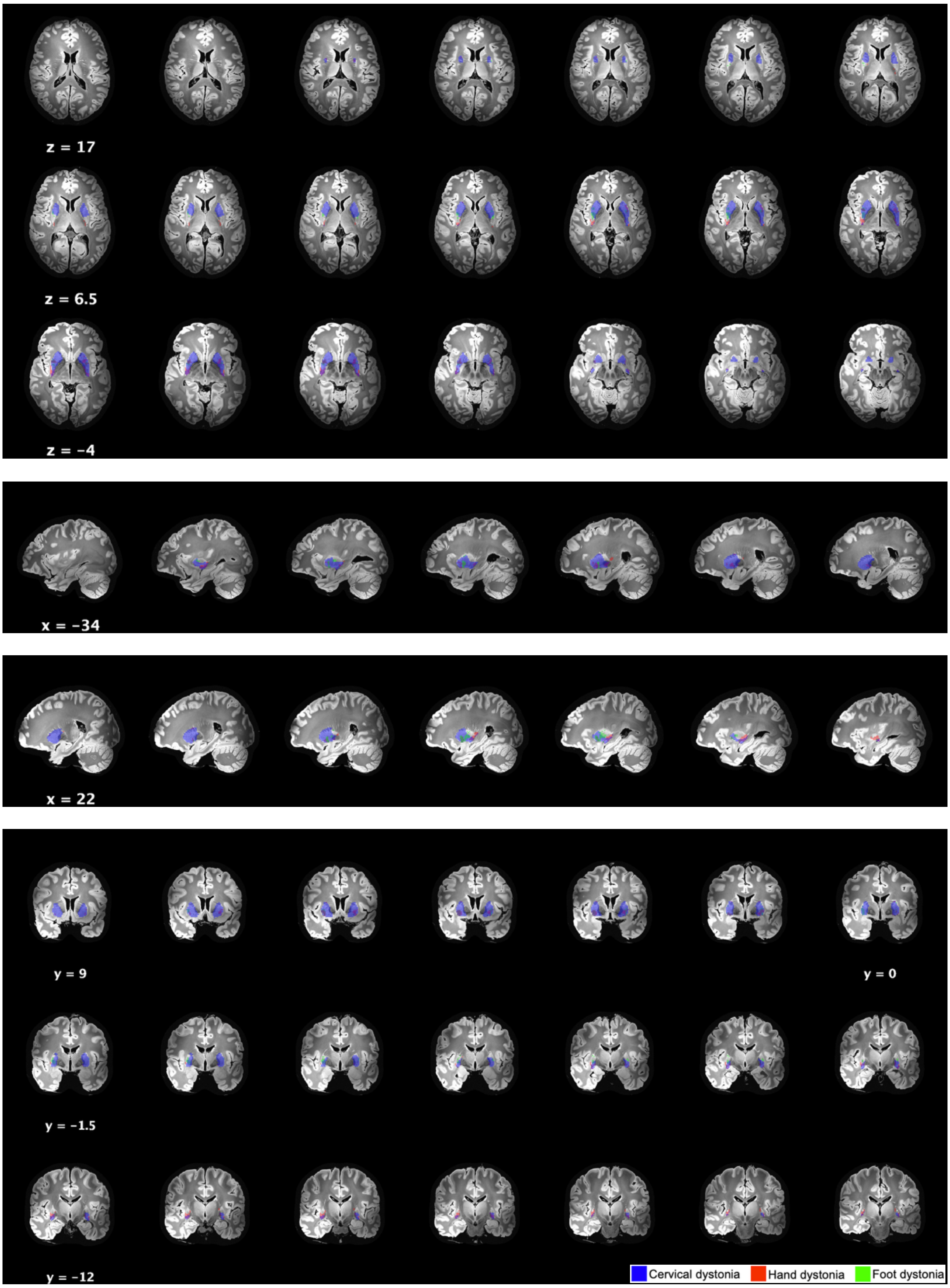

**Supplementary File 13C.** Additional slices of cervical, hand, and foot dystonia lesion networks in the putamen.
